# Association between late-life air pollution exposure and medial temporal lobe atrophy in older women

**DOI:** 10.1101/2023.11.28.23298708

**Authors:** Xinhui Wang, Lauren E. Salminen, Andrew J. Petkus, Ira Driscoll, Joshua Millstein, Daniel P. Beavers, Mark A. Espeland, Guray Erus, Meredith N. Braskie, Paul M. Thompson, Margaret Gatz, Helena C. Chui, Susan M Resnick, Joel D. Kaufman, Stephen R. Rapp, Sally Shumaker, Mark Brown, Diana Younan, Jiu-Chiuan Chen

**Affiliations:** Department of Neurology, University of Southern California, Los Angeles, California; Mark and Mary Stevens Neuroimaging and Informatics Institute, Keck School of Medicine, University of Southern California, Los Angeles, California; Department of Medicine, University of Wisconsin-Madison, Madison, Wisconsin; Department of Population and Public Health Sciences, University of Southern California, Los Angeles, California; Departments of Statistical Sciences, Wake Forest University, Winston-Salem, North Carolina; Department of Internal Medicine, Wake Forest School of Medicine, Winston-Salem, North Carolina; Department of Biostatistics and Data Sciences, Wake Forest School of Medicine, Winston-Salem, North Carolina; Center for Biomedical Image Computing and Analytics (CBICA), University of Pennsylvania, Philadelphia, Pennsylvania; Center for Economic and Social Research, University of Southern California, Los Angeles, California; The Laboratory of Behavioral Neuroscience, National Institute on Aging, Baltimore, Maryland; Departments of Environmental & Occupational Health Sciences, Medicine (General Internal Medicine), and Epidemiology, University of Washington, Seattle, Washington; Departments of Psychiatry and Behavioral Medicine, Wake Forest School of Medicine, Winston-Salem, North Carolina; Department of Social Sciences and Health Policy, Wake Forest School of Medicine, Winston-Salem, North Carolina

**Keywords:** air pollution, medial temporal lobe, atrophy, AD neuropathology, parahippocampal gyrus, entorhinal cortex

## Abstract

**Background:** Ambient air pollution exposures increase risk for Alzheimer’s disease (AD) and related dementias, possibly due to structural changes in the medial temporal lobe (MTL). However, existing MRI studies examining exposure effects on the MTL were cross-sectional and focused on the hippocampus, yielding mixed results.

**Method:** To determine whether air pollution exposures were associated with MTL atrophy over time, we conducted a longitudinal study including 653 cognitively unimpaired community-dwelling older women from the Women’s Health Initiative Memory Study with two MRI brain scans (MRI-1: 2005-6; MRI-2: 2009-10; M_age_ at MRI-1=77.3±3.5years). Using regionalized universal kriging models, exposures at residential locations were estimated as 3-year annual averages of fine particulate matter (PM_2.5_) and nitrogen dioxide (NO_2_) prior to MRI-1. Bilateral gray matter volumes of the hippocampus, amygdala, parahippocampal gyrus (PHG), and entorhinal cortex (ERC) were summed to operationalize the MTL. We used linear regressions to estimate exposure effects on 5-year volume changes in the MTL and its subregions, adjusting for intracranial volume, sociodemographic, lifestyle, and clinical characteristics.

**Results:** On average, MTL volume decreased by 0.53±1.00cm^3^ over 5 years. For each interquartile increase of PM_2.5_ (3.26µg/m^3^) and NO_2_ (6.77ppb), adjusted MTL volume had greater shrinkage by 0.32cm^3^ (95%CI=[-0.43,-0.21]) and 0.12cm^3^ (95%CI=[-0.22,-0.01]), respectively. The exposure effects did not differ by *APOE* ε4 genotype, sociodemographic, and cardiovascular risk factors, and remained among women with low-level PM_2.5_ exposure. Greater PHG atrophy was associated with higher PM_2.5_ (b=-0.24, 95%CI=[-0.29,-0.19]) and NO_2_ exposures (b=-0.09, 95%CI=[-0.14,-0.04]). Higher exposure to PM_2.5_ but not NO_2_ was also associated with greater ERC atrophy. Exposures were not associated with amygdala or hippocampal atrophy.

**Conclusion:** In summary, higher late-life PM_2.5_ and NO_2_ exposures were associated with greater MTL atrophy over time in cognitively unimpaired older women. The PHG and ERC - the MTL cortical subregions where AD neuropathologies likely begin, may be preferentially vulnerable to air pollution neurotoxicity.

**Highlights:**

- First longitudinal study on air pollution and medial temporal lobe (MTL) volume.
- Late-life PM_2.5_ and NO_2_ associated with MTL atrophy over time in older women.
- Heterogeneous adverse effects were observed across different subregions of the MTL.
- Results not differ by *APOE* genotype, age, education, or cardiovascular risk factors.
- Adverse effects remained at low-level exposure compliant with regulatory standards.

## 1. Introduction

Alzheimer’s disease (AD) and related dementias (ADRD) are leading causes of death and disability worldwide, disproportionately affecting women over age 65.^1^ The progression of ADRD occurs along a continuum of neuropathological processes and brain atrophy that eventually leads to cognitive decline and impaired activities of daily living.^2,3^ Early AD is characterized by changes in memory-related processes that are subserved by the medial temporal lobe (MTL), including the hippocampus, amygdala, parahippocampal gyrus (PHG), and entorhinal cortex (ERC).^4–8^ Cumulative evidence has shown that late-life exposure to ambient air pollution, including fine particulate matter (PM_2.5_) and traffic-related pollution (NO_2_), is a risk factor for AD-related neuropathology,^9,10^ memory-related cognitive decline, and ADRD.^11–13^ However, brain MRI studies linking air pollution neurotoxicity to MTL atrophy in humans are less conclusive, especially at the preclinical stage, partly due to cross-sectional designs that yielded mixed results. No studies have investigated the neurotoxic effects of exposures in MTL subregions either.^10,14–19^ Additionally, it remains unclear whether established risk factors for late-onset AD, such as cardiovascular disease (CVD) and the Apolipoprotein E (*APOE*) genotype, modulate the neurotoxic effects of air pollution on AD-related neurodegeneration.^20–25^ To address these knowledge gaps, we conducted a longitudinal study to assess whether PM_2.5_ and NO_2_ contribute to atrophy of the MTL and its subregions in cognitively unimpaired older women, and evaluated whether associations differ by population characteristics and AD risk factors.

## 2. Methods

### 2.1 Participants and study design

We examined community-dwelling women enrolled in the Women’s Health Initiative (WHI) Memory Study (WHIMS)^26^ who underwent two MRI scans.^27,28^ WHIMS is an ancillary study of WHI – hormone therapy trials, designed to investigate postmenopausal hormone therapy on cognitive function and dementia risk. Women were ≥65 years of age and free of dementia at enrollment. Between April 2005 and January 2006, 1405 women completed a baseline MRI scan (MRI-1). Approximately half of these women (n=720) completed a follow-up MRI scan (MRI-2) in 2009-2010, with average 4.7 years of follow-up. Since this study focused on examining air pollution neurotoxicity in the preclinical stage, we excluded 11 women with mild cognitive impairment (MCI) or probable dementia at MRI-1 and 56 women missing key covariates, rendering a final analytic sample of 653 cognitively unimpaired women (Figure 1).

**Figure 1.**
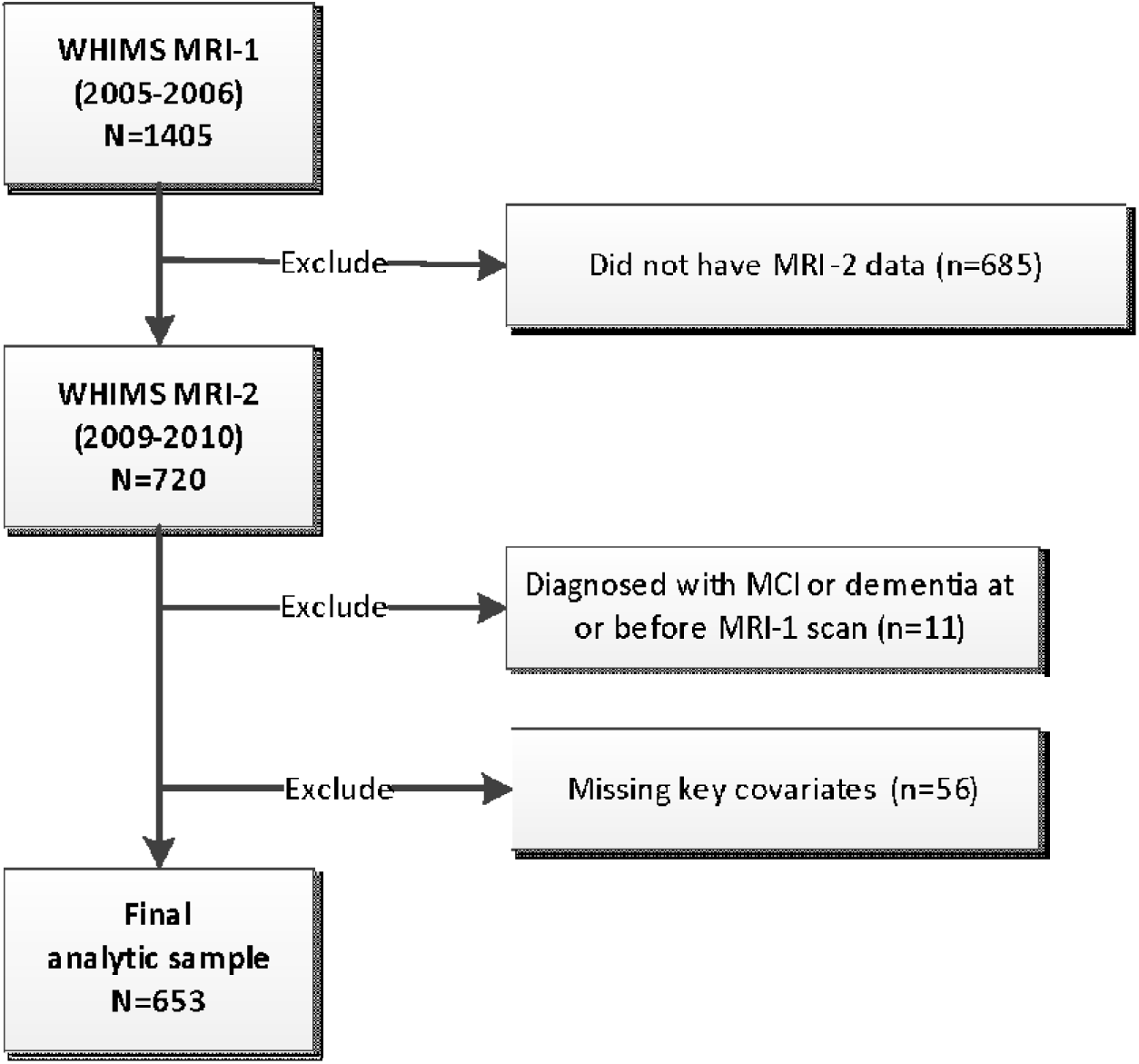
Flowchart of study population. Abbreviations: MCI, mild cognitive impairment; MRI, magnetic resonance imaging; WHIMS, Women’s Health Initiative Memory Study.

Study protocols were approved by the Institutional Review Board at the University of Southern California. Written informed consent was obtained from all participants as part of the WHIMS-MRI study.

### 2.2 Air pollution exposure estimation

Participants’ residential addresses were prospectively collected at each WHI assessment since WHI inception in 1993 and updated at least semiannually during regular follow-up contacts or by participant reporting change of address between regular contacts.^29^ At each geocoded residential location, annual mean concentrations of ambient PM_2.5_ in μg/m^3^ and NO_2_ in ppb were estimated using validated regionalized national universal kriging models with partial least squares regression of geographic covariates and US Environmental Protection Agency monitoring data. Over 300 geographic covariates covering categories of population, land use, vegetative index, impervious surfaces, roadway, and proximity to features were used in the national models for PM_2.5_ estimation.^30^ For NO_2_ estimation, over 400 geographic covariates covering proximity and buffer measures as well as satellite data were used in prediction.^31^ The average cross-validation R^2^ was 0.88 for PM_2.5_ and 0.85 for NO_2_. We used the annual estimates of each pollutant to calculate the average spanning the 3-year time window prior to MRI-1 with the length of stay at each residential location within the 3 years as the weight, accounting for residential mobility.

### 2.3 MRI acquisition and processing

MRI scans were collected on 1.5T scanners at 14 WHI centers using standardized acquisition protocols developed by the WHIMS-MRI Quality Control Center at the University of Pennsylvania, Philadelphia.^32–34^ The scan series for volumetric imaging used a 22cm field of view and a 256×256 acquisition matrix and the following pulse sequences: oblique axial spin density/T2-weighted spin echo images, oblique axial fast fluid-attenuated inversion recovery (FLAIR) T2-weighted spin echo image and oblique axial fast spoiled 3D T1-weighted gradient echo images. Pulse sequence parameters are provided in earlier work.^32,33^ Trained technicians at each site completed rigorous quality control procedures (e.g., magnetic field homogeneity evaluation, slice thickness and position accuracy, RF coil checks, etc.) outlined by the WHIMS-MRI program, which was adapted from the American College of Radiology.

Regions of interest (ROIs) were extracted using a multi-atlas region segmentation (MUSE) that transforms region-specific labeled atlases into a harmonized map. Specifically, MUSE follows a voxel-based spatial adaptation strategy to transform multiple atlases with different warping algorithms and regularization parameters into an ensemble-based parcellation of anatomical reference labels. This approach leads to robust segmentation accuracy and is superior to other multi-atlas segmentation and label fusion methods, especially for multi-site MRI investigations and longitudinal analyses.^35,36^

Gray matter volumes of the hippocampus, amygdala, PHG, and ERC were estimated and summed across left and right hemispheres. Total MTL volume was operationally defined as the summed volumes of these four bilateral ROIs. MTL atrophy was quantified as the difference between volumes measured at MRI-1 and MRI-2, divided by the years between the two MRI scans and then multiplied by 5 to represent 5-year volume changes.

### 2.4 Covariates of interest

At WHI inception (1993–1998), structured questionnaires were administered to gather information on age, race/ethnicity, socioeconomic factors (education, family income, and employment status), and lifestyle factors (smoking status, alcohol use, and physical activity). Except for continuous age at MRI-1, other variables with categorical levels reported in tables were used in analyses. As this was a predominantly White sample, the race/ethnicity variable was coded as: “White (not Hispanic)” vs. “Other ethnic or racial background” which included “Hispanic/Latino”, “American Indian/Alaska Native”, “Asian”, “Black”, or “More than one race”.

Clinical characteristics collected at WHI inception included body mass index (BMI, calculated using measured weight and height), depressive symptoms using the Center for Epidemiologic Studies Depression Scale short form,^37^ self-reported prior use of postmenopausal hormones, and self-reported history of CVD (e.g., heart problems, problems with blood circulation, blood clots) and related risk factors (e.g., hypertension, hypercholesterolemia or diabetes mellitus) which were validated previously.^38–40^ Because few women endorsing a history of diabetes (n=16) or hypercholesterolemia (n=97), we created a binary variable to indicate if a person had none or at least one type of CVD risk factors to boost statistical power.

Besides information collected at WHI inception, lifestyle factors, BMI, hypertension, and CVD history were also updated at MRI-1. Contextual socioeconomic characteristics of residential neighborhoods (e.g., neighborhood socioeconomic status, nSES) were measured at both WHI inception and MRI-1 using US Census tract-level data.^41^ Higher nSES scores indicated more socioeconomically-favorable neighborhoods. *APOE* genotype (ε3/ε3, ε3/ε4, or ε4/ε4) was measured in a subset of 522 women. Details are available in the Supplement.

### 2.5 Statistical Approach

We used linear regressions to estimate the associations between baseline air pollution exposures and 5-year volume changes in the total MTL and its subregions, adjusting for intracranial volume (ICV), sociodemographic variables (age, geographical region, race/ethnicity, education, income, employment status, and nSES), lifestyle factors (smoking, alcohol use, and physical activity), prior postmenopausal hormone use, WHI hormone therapy assignment, and clinical characteristics (BMI, depressive symptoms, CVD risk factors) collected at WHI inception gradually in main analyses (Table 2). To account for attrition from MRI-1 to MRI-2, we incorporated inverse-probability weighting in all models. We conducted moderation analyses to determine whether exposure associations with MTL atrophy differed by age, education, CVD risk factors, BMI, and *APOE* ε4 genotype (carriers vs. non-carriers). To put the estimated associations in context, we conducted an *ad hoc* Cox proportional hazard regression in women without dementia before MRI-1 to estimate how exposure-related MTL atrophy would translate to dementia risk, using outcome data from the WHIMS protocols. Details are described in the Supplement.

Additional analyses were conducted to evaluate the robustness of our findings. First, we restricted our analyses to women with PM_2.5_ exposure below the NAAQS (12 μg/m^3^) to evaluate low-level exposure effects. Second, we refit the models using covariates updated at MRI-1 to address residual confounding due to temporal misspecification of potential confounders (lifestyle factors, nSES, and clinical variables). Third, we refit the models with further adjusting for the corresponding MRI-1 volume to assess whether the regression to the mean in MRI volume changes may impact the observed associations. Lastly, we excluded women who developed MCI or dementia between MRI-1 and MRI-2 to determine whether observed associations were driven by underlying dementia risk.

All statistical analyses were performed using R 4.1.2 and SAS 9.4. Statistical significance for inferential analyses were interpreted at the 0.05 alpha level. Multiple comparison correction was done for analyses on the four MTL subregions using the Benjamini–Hochberg false discovery rate (FDR) approach.^42^

## 3. Results

### 3.1 Sample characteristics

Compared to the 752 women excluded from this study (Figure 1), women in the analytic sample (n=653) tended to be younger, had lower exposures and larger MTL volumes, and had more overweight (BMI in 25-29 kg/m^2^) at MRI-1. A greater proportion identified as non-Hispanic White and had higher family income but had fewer CVD risk factors (Tables S1&S2).

Non-Hispanic White women had significantly lower PM_2.5_ and NO_2_ exposures at MRI-1 than those of other ethnic and racial backgrounds (Table 1). Women living in the West had lower PM_2.5_ exposures while women living in the South had much lower NO_2_ exposures than those from other regions. Compared to their counterparts, women without prior use of postmenopausal hormones or those living in the most socioeconomically-*un*favorable neighborhoods had higher PM_2.5_ exposures. By contrast, NO_2_ exposures were the highest in the most socioeconomically-favorable neighborhoods, and in women aged ≥80 years, completed ≥4 years of college, currently smoking, past drinkers or those drinking <1 drink/day at MRI-1. On average, total MTL volume decreased by 0.53±1.00 cm^3^ over 5 years and was more pronounced in women aged ≥80 years, lived in the Northeast, or with BMI <25 kg/m^2^ at MRI-1 (Table 1).

**Table 1.**
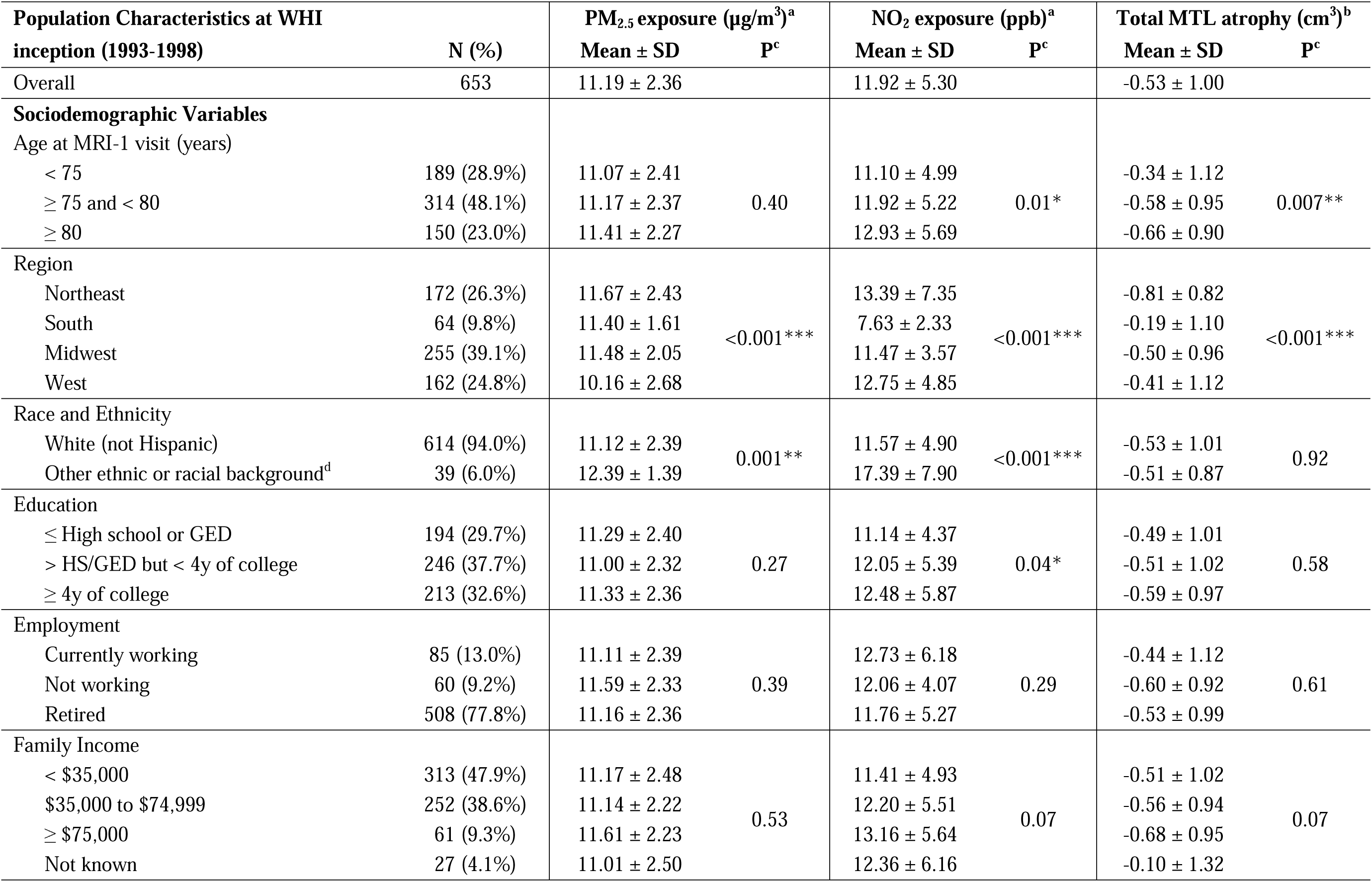
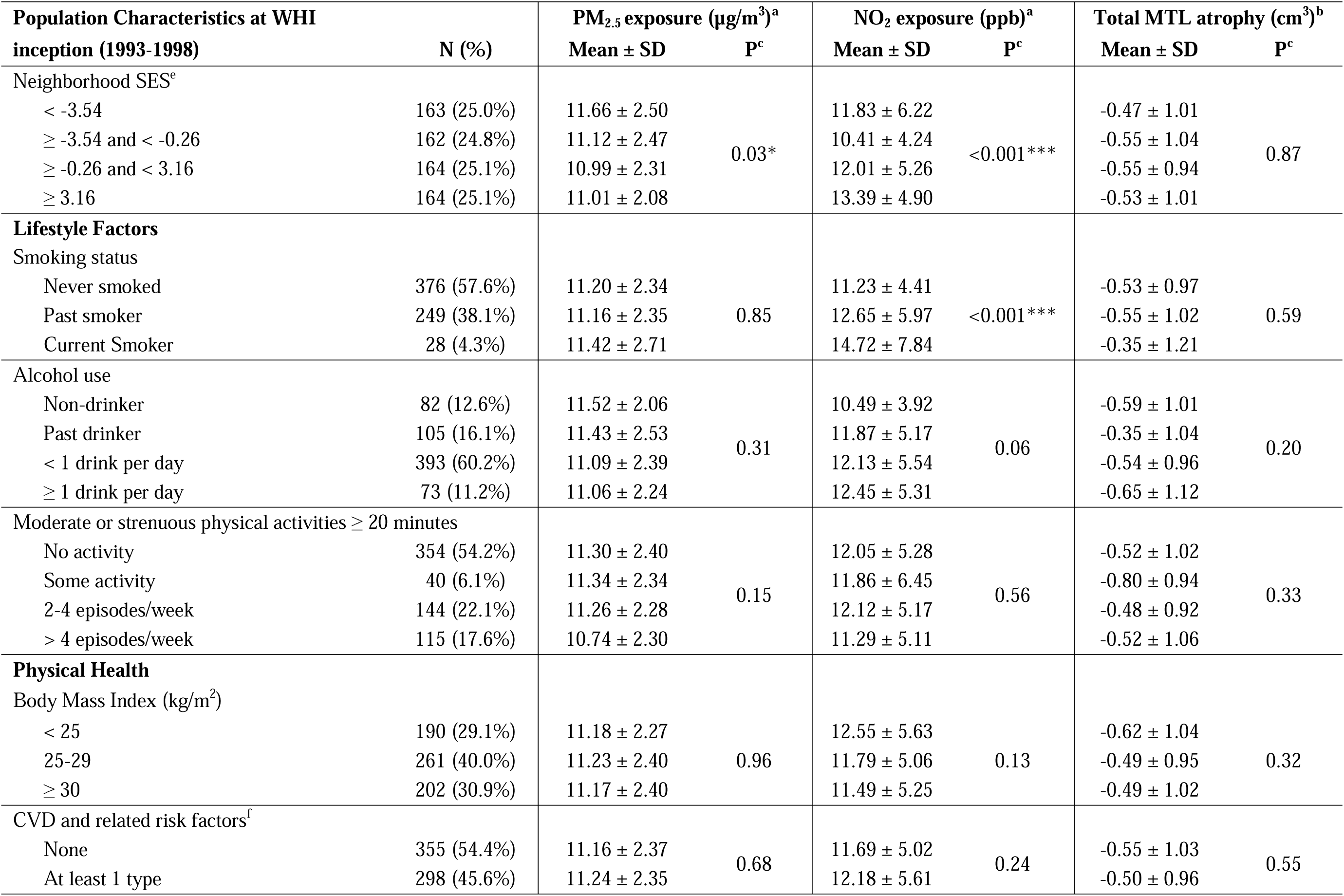
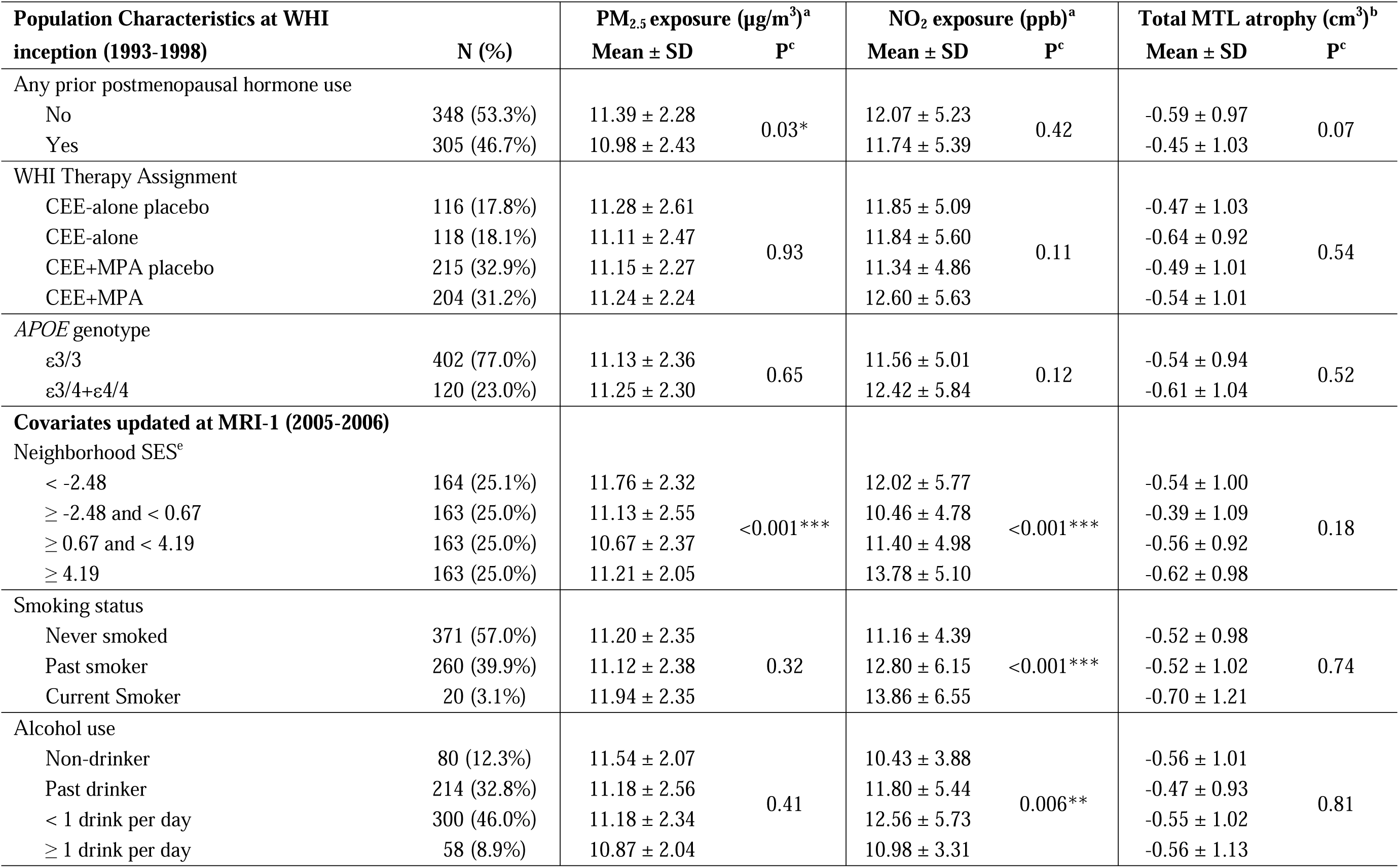
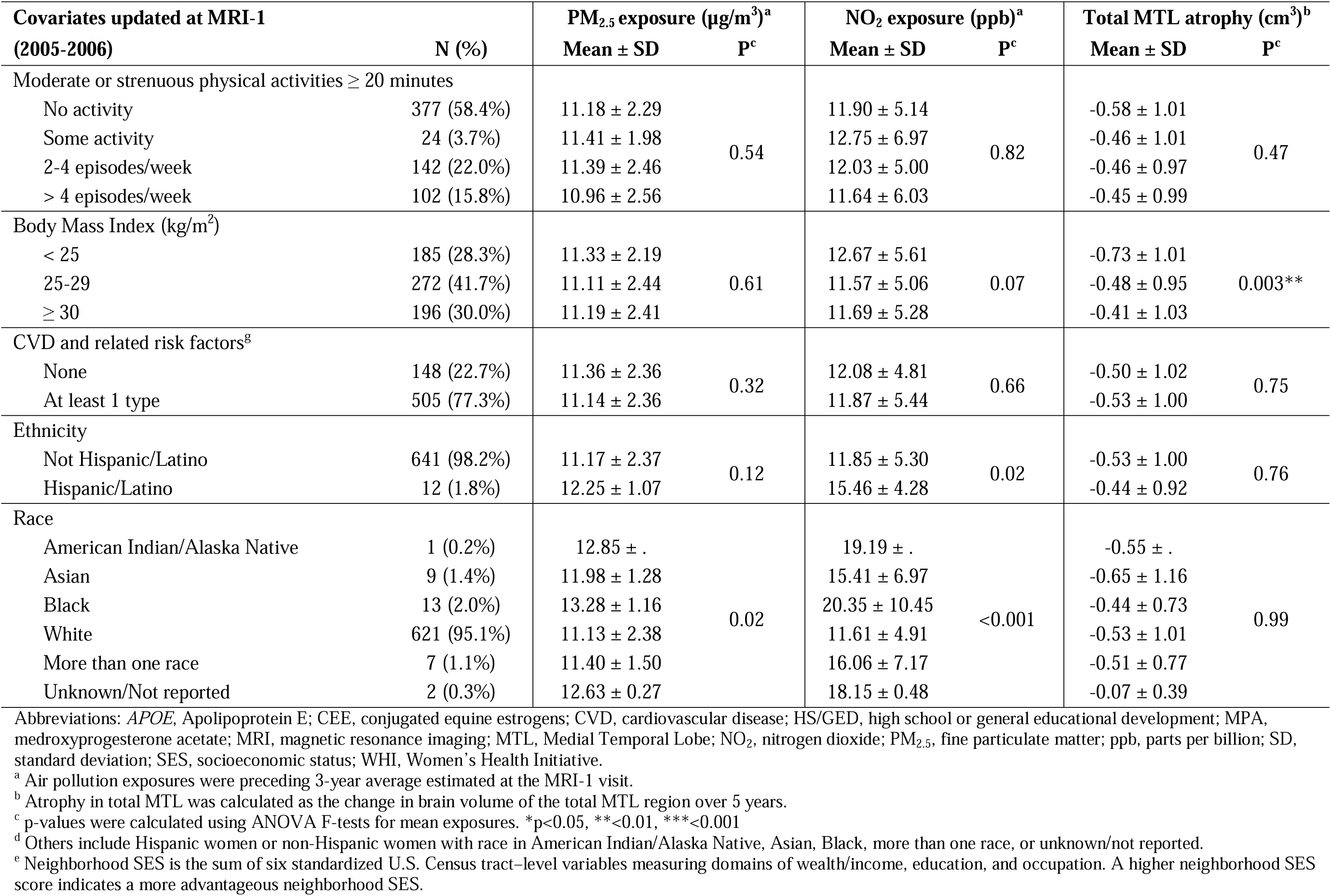
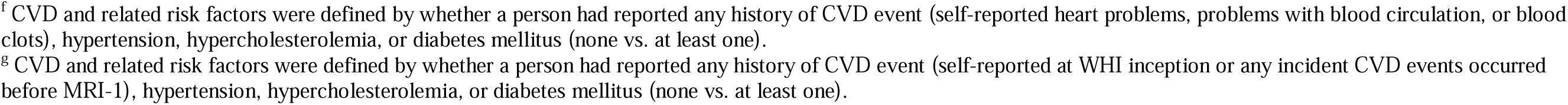
Distribution of air pollution exposures and total medial temporal lobe atrophy by population characteristics.

### 3.2 Associations between exposures and MTL atrophy

Table 2 shows the associations between exposures and MTL atrophy across multiple models adjusting for different covariates. For each interquartile range (IQR) increase in PM_2.5_ (IQR=3.26 µg/m^3^) and NO_2_ (IQR=6.77 ppb), MTL volume decreased by 0.32 cm^3^ (95% CI=[−0.43, −0.21], p<0.01) and 0.12 cm^3^ (95% CI=[−0.22, −0.01], p=0.03) over 5 years, adjusting for ICV, age, geographic region, race/ethnicity, SES measures, lifestyle factors, and clinical characteristics (Model E, Table 2). The observed exposure-related MTL atrophies were equivalent to 17% (95% CI=[11%, 24%]) and 6% (95% CI=[0.6%, 12%]) increased dementia risk, respectively.

**Table 2.** Associations between air pollution exposures and medial temporal lobe atrophy.

| Model | PM<sub>2.5</sub> exposure<sup>a</sup> |  |  | NO<sub>2</sub> exposure<sup>a</sup> |  |  |
| --- | --- | --- | --- | --- | --- | --- |
|  | b<sup>b</sup> | 95% CI | p | b<sup>b</sup> | 95% CI | p |
| Model A | -0.32 | -0.42, -0.21 | <0.01 | -0.16 | -0.25, -0.07 | <0.01 |
| Model B | -0.31 | -0.42, -0.20 | <0.01 | -0.11 | -0.22, -0.01 | 0.03 |
| Model C | -0.32 | -0.43, -0.21 | <0.01 | -0.12 | -0.22, -0.01 | 0.03 |
| Model D | -0.32 | -0.43, -0.21 | <0.01 | -0.11 | -0.22, -0.01 | 0.03 |
| Model E | -0.32 | -0.43, -0.21 | <0.01 | -0.12 | -0.22, -0.01 | 0.03 |
| Model F | -0.31 | -0.42, -0.20 | <0.01 | -0.11 | -0.21, -0.002 | 0.047 |
| Model G | -0.32 | -0.43, -0.21 | <0.01 | -0.12 | -0.22, -0.01 | 0.03 |

Abbreviations: CI: confidence interval; MRI, magnetic resonance imaging; NO<sub>2</sub>, nitrogen dioxide; PM<sub>2.5</sub>, fine particulate matter; ppb, parts per billion; SES, socioeconomic status; WHI, Women's Health Initiative.
<sup>a</sup> Air pollution exposures were preceding 3-year average estimated at the MRI-1 visit.
interquartile range (IQR)<sub>PM2.5</sub> = 3.26 µg/m<sup>3</sup>; IQR<sub>NO2</sub> = 6.77 ppb
<sup>b</sup> b represents the average change in brain volume (cm<sup>3</sup>) over 5 years for each IQR increase of 3-year average exposure. A negative b means higher air pollution exposure was associated with a greater atrophy in medial temporal lobe over 5 years.
Model A: incorporated inverse-probability weighting approach and adjusted for the intracranial volume and age
Model B: adjusted for Model A covariates + geographic region, race/ethnicity, education, income, employment status, and neighborhood SES
Model C: adjusted for Model B covariates + lifestyle factors (smoking; alcohol use; physical activities), prior postmenopausal hormone use, and WHI hormone therapy assignment
Model D: adjusted for Model C covariates + body mass index and depressive symptoms (logit transformation of the raw score)
Model E: adjusted for Model D covariates + cardiovascular disease and related risk factors
Model F: adjusted for Model E covariates with lifestyle factors, body mass index, neighborhood SES, and cardiovascular disease and related risk factors updated at MRI-1 visit.
Model G: adjusted for Model E covariates + medial temporal lobe volume at MRI-1 visit.

Associations between exposure and total MTL atrophy did not change much when partial covariates were removed (Models B - D vs. E) and remained significant with adjustment of covariates updated at MRI-1 (Model F) or MTL volume at MRI-1 (Model G, Table 2). When restricting the analyses to individuals with low-level exposures (PM_2.5_<12 μg/m^3^), the associations became stronger (Table S5). After excluding 33 women with incident MCI or dementia before MRI-2, the estimated associations did not change for PM_2.5_ and became stronger for NO_2_ (b=-0.14, 95% CI=[−0.25, −0.04], p<0.01; Model E, Table S6). We found no statistical evidence that the observed adverse exposure effects on MTL atrophy differed by age, education, BMI, CVD risk factors, or *APOE* genotype (Figure 2).

**Figure 2.**
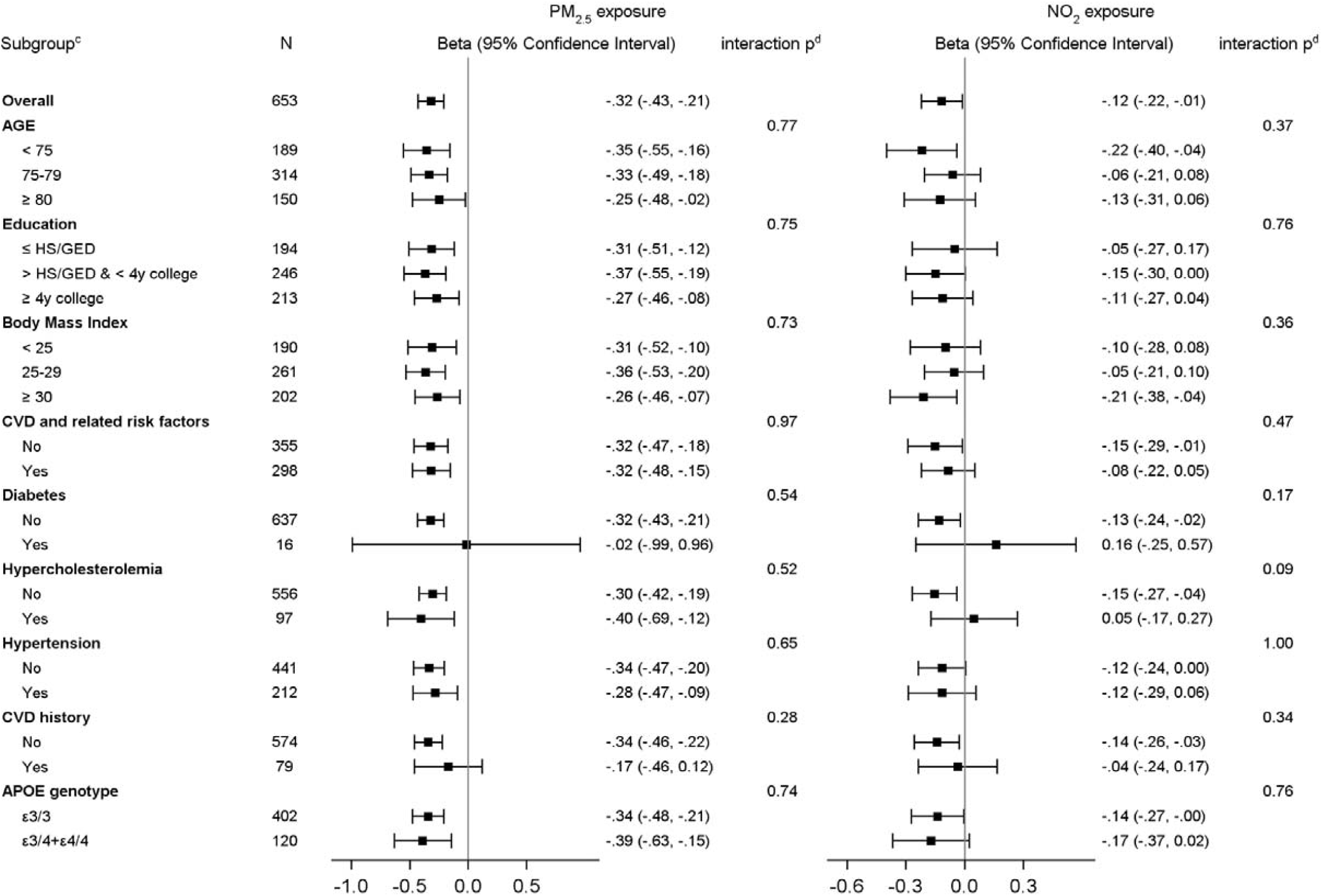
Estimated associations^a^ between air pollution exposures^b^ and atrophy in the medial temporal lobe, stratified by population characteristics. Abbreviations: *APOE*, Apolipoprotein E; CVD, cardiovascular disease; HS/GED, high school/general educational development; IQR, interquartile range; MRI, magnetic resonance imaging; NO_2_, nitrogen dioxide; PM_2.5_, fine particulate matter; ppb, parts per billion; WHI, Women’s Health Initiative. Legend: The bars and whisker represent the regression coefficients and corresponding 95% confidence intervals. ^a^ Associations were estimated as volume change (in cm^3^) in medial temporal lobe over 5 years for each IQR increase of air pollution exposure (IQR_PM2.5_ = 3.26 μg/m^3^; IQR_NO2_ = 6.77 ppb), incorporating inverse-probability weighting approach and adjusting for intracranial volume, geographic region, age, race/ ethnicity, education, income, employment status, neighborhood socioeconomic status, lifestyle factors (smoking, drinking, and physical activities), prior postmenopausal hormone use, WHI hormone therapy assignment, body mass index, depressive symptoms, and CVD and related risk factors. The results of overall samples were from Model E of Table 2. ^b^ Air pollution exposures were preceding 3-year average estimated at the MRI-1 visit. ^c^ CVD and related risk factors were defined by whether a person had reported any history of CVD event (self-reported heart problems, problems with blood circulation, or blood clots), hypertension, hypercholesterolemia, or diabetes mellitus (none vs. at least one). ^d^ p-Value was calculated using Wald t test for the interaction between exposure and each subgroup unadjusted for multiple comparison. After controlling for multiple comparison using Benjamini–Hochberg approach, false discovery rate corrected p-values > 0.05 for all interaction tests.

Exposure-related brain atrophy differed across MTL subregions (Figure 3, Table S4). For each respective IQR increase of PM_2.5_ and NO_2_, PHG volume declined by 0.24 cm^3^ (95% CI=[−0.29, −0.19], FDR-corrected p<0.01) and 0.09 cm^3^ (95% CI=[−0.14, −0.04], FDR-corrected p<0.01) over 5 years, after adjusting for all potential confounders (Model E, Table S4). The corresponding ERC volume declined by 0.06 cm^3^ over 5 years (95% CI=[−0.10, −0.01], FDR-corrected p=0.04) per IQR increase of PM_2.5_, but not with NO_2_. Neither pollutant was associated with amygdala or hippocampal atrophy (Table S4).

**Figure 3.**
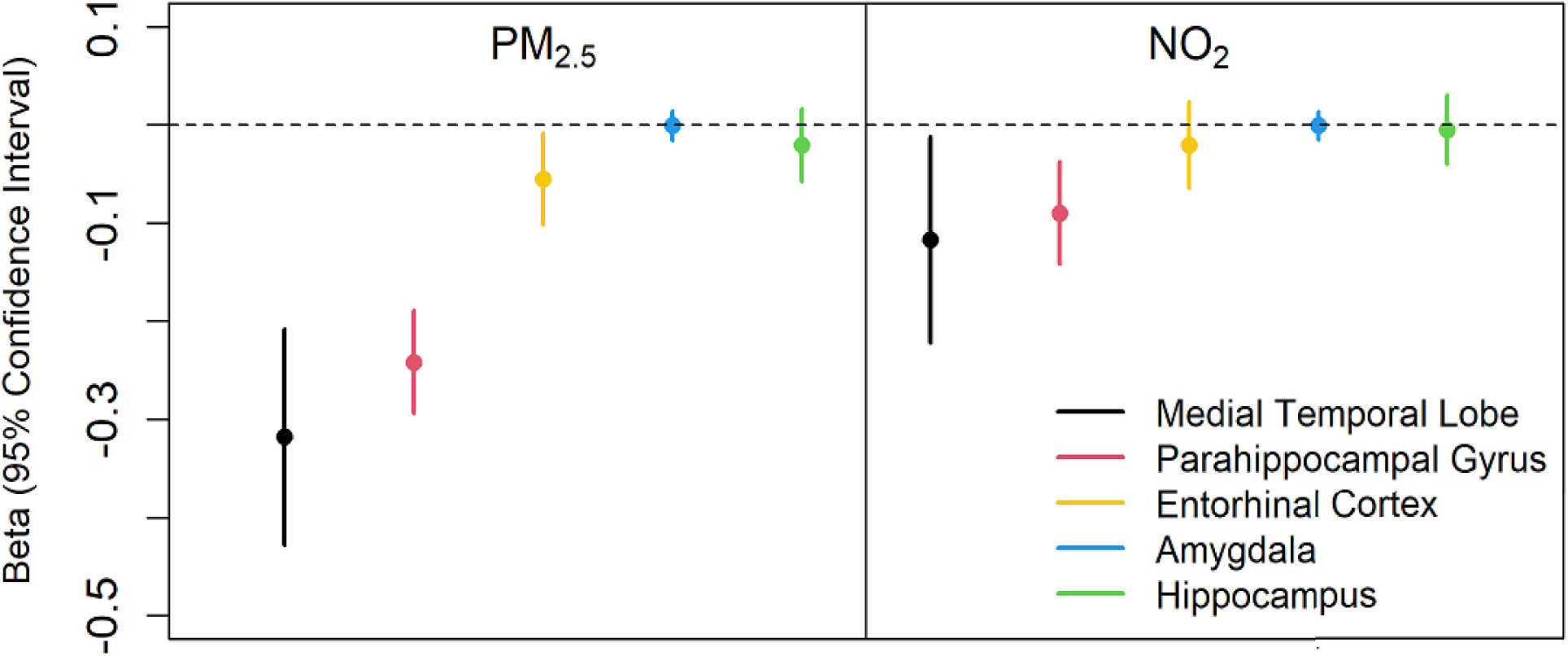
Estimated associations between air pollution exposures and atrophy in the medial temporal lobe and its subregions. Abbreviations: IQR, interquartile range; MRI, magnetic resonance imaging; NO_2_, nitrogen dioxide; PM_2.5_, fine particulate matter; ppb, parts per billion; WHI, Women’s Health Initiative. Legend: The dots and whiskers represent the regression coefficients and corresponding 95% confidence intervals. The regression coefficients were estimated as volume changes (in cm^3^) over 5 years for each IQR increase of air pollution exposure (IQR_PM2.5_ = 3.26 µg/m^3^; IQR_NO2_ = 6.77 ppb). Results illustrated in this figure were from Model E of Table 2 and Table S4, which incorporated inverse-probability weighting approach and adjusted for intracranial volume, geographic region, age, race/ethnicity, education, income, employment status, neighborhood socioeconomic status, lifestyle factors (smoking, drinking, and physical activities), prior postmenopausal hormone use, WHI hormone therapy assignment, body mass index, depressive symptoms, and cardiovascular disease and related risk factors.

## 4. Discussion

This is the first longitudinal study linking late-life air pollution exposures to preclinical MTL atrophy in community-dwelling cognitively unimpaired older women. We found that women living in locations with higher levels of PM_2.5_ and NO_2_ had greater MTL atrophy over time, and these associations could not be explained by sociodemographic, lifestyle, and clinical characteristics. Exposure-related brain atrophy varied across MTL subregions, with significant adverse effects observed for the PHG and ERC, but not the amygdala or hippocampus. These associations persisted in women who remained cognitively unimpaired over the study period and also in women living in locations with PM_2.5_ below the current EPA standard. Exposure-related MTL atrophy did not differ substantially by sociodemographic variables, cardiovascular risk factors, or *APOE* genotype. Collectively these results support the contribution of air pollution neurotoxicity on preclinical MTL neurodegeneration in older women.

Previous MRI studies investigating exposure effects on the brain regions that are vulnerable to AD in older adults have examined the hippocampus^10,14–19^ or “AD signature” areas.^17,21,43^ They were cross-sectional and largely reported no associations. Longitudinal studies offer a better understanding of how air pollution exposures influence intraindividual brain changes. Using the WHIMS-MRI data, our group published the first longitudinal MRI study linking PM_2.5_ exposure to the spatial extent of AD-related neurodegeneration^43^, as reflected by the AD pattern similarity score, which summarizes multiple areas of brain atrophy beyond the MTL, but does not allow the examination on specific regions. Because MTL regions contribute critically to memory-related processes and are more susceptible to early AD-related brain disruption,^6,44^ focusing on MTL atrophy could provide better insight on air pollution neurotoxicity on brain aging at preclinical stage of AD progression. The results presented herein add to the literature by showing adverse longitudinal effects of both PM_2.5_ and NO_2_ on progressive MTL atrophy in cognitively unimpaired older women, specifically in cortical MTL subregions.

Associations between long-term air pollution exposure and hippocampal volume in humans are complex. A recent meta-analysis identified a significant PM_2.5_ exposure effect on the adult hippocampus,^45^ yet three studies that reported null associations were excluded.^14–16^ In addition, even though null associations were found on the whole hippocampus volume in three studies included in the meta-analyses,^17,18,21^ non-independent effect size estimates ^46^ that were modeled separately by sex,^18^ geographic location,^17^ or on left and right hippocampi were used in the meta analyses.^19,21^ Thus, the observed lack of exposure effect on hippocampal shrinkage in our study is consistent with 6 out of 7 cross-sectional studies that reported no association between PM_2.5_ and whole hippocampal volume in older adults.^14–19, 21^

To our knowledge only one cross-sectional study has examined the associations of air pollution exposure on MTL subregions beyond the hippocampus.^18^ Specifically, Cho et al.^18^ reported inverse associations between PM_2.5_ (1-year average) and NO_2_ (5-year average) exposure and cortical thickness in the PHG, but not the ERC in cognitively unimpaired older adults living in South Korea. The authors also reported a significant association between higher NO_2_ (but not PM_2.5_) and smaller amygdala volume. Our longitudinal results further demonstrate that cortical atrophy in the PHG, and to some extent the ERC, are important MTL targets of air pollution neurotoxicity at the preclinical stage. We did not observe any longitudinal effect of exposures on subcortical MTL volumes (hippocampus and amygdala).

Adverse exposure associations with cortical (versus subcortical) MTL atrophy is consistent with the established pattern of brain atrophy in preclinical AD that typically begins in the cortex and later impacts subcortical areas.^44,47^ Further, PHG and ERC atrophy has been shown to predict incident dementia risk in longitudinal human MRI studies, whereas hippocampal atrophy does not predict dementia risk after controlling for ERC volume.^48^ Exposure-related atrophy in the PHG and ERC has important functional implications, as both structures are involved in information encoding, consolidation, and retrieval as well as spatiotemporal memory.^49–51^ Indeed, atrophy in these structures may explain previously reported associations between air pollution exposure and cognitive difficulties on tests of memory.^52–55^ These structures are also important for olfactory processing functions that mediate memory formation,^56,57^ are associated with air pollution exposure,^58,59^ and are among the earliest to shrink in preclinical ADRD.^60,61^

How air pollution neurotoxicity affects neuropathological processes along the ADRD continuum at the preclinical stage remains largely unknown. Animal studies have shown that air pollution exposure contributes to AD pathogenic processes such as beta-amyloid (Aβ) and tau deposition,^62^ and this was supported in recent human studies using PET imaging ^9^, CSF and plasma biomarkers.^10,63^ Progressive exposure-related MTL atrophy may also result from neuropathological processes apart from preclinical AD,^6^ such as hippocampal sclerosis,^64^ primary age-related tauopathy (PART),^65^ and limbic-predominant age-related TDP-43 encephalopathy (LATE).^66^ Although the neuropathological processes underlying the observed associations in our study are unclear, our findings should motivate future studies that use PET and other biomarkers to test the hypothesized mediating role of Aβ or tau between exposures and MTL atrophy across different age-related neurodegenerative conditions.

Our study has some limitations. First, we cannot make inferences about possible effects of exposure during midlife, which may also be a vulnerability period for the preclinical AD pathogenic processes.^67,68^ Second, we could not rule out the possibility of unmeasured confounding by other environmental factors (such as, noise and green space). However, it is noteworthy that scientific evidence for the early neurodegeneration affected by these other environmental exposures is sparse. Third, prior data from animal models suggest that exposure may selectively affect the CA1 subfield of the hippocampus,^69^ yet our study did not include fine segmentation of hippocampal subfields and this deserves further examination in humans. Fourth, the MUSE MRI protocols do not parcellate local white matter, but investigations of white matter microstructure may inform the heterogeneous associations between exposures and MTL subregions. Lastly, our findings cannot be generalized to men or younger women.

Major strengths of this study include our longitudinal design and rigorous analytic approach that allowed us to evaluate intraindividual changes in exposure-related brain outcomes. Additionally, the high-quality, comprehensive data of the WHIMS cohort allowed us to adjust for a variety of covariates and evaluate potential impacts related to the development of incident dementia. Finally, the MUSE MRI protocol used herein was designed to improve multi-site registration issues for longitudinal analyses, as it fuses multiple warped atlases that are not site-specific harmonized to define the ROIs.^35^

## 5. Conclusions

In cognitively unimpaired older women, late-life exposures to outdoor air pollution contribute to preclinical cortical atrophy of MTL, including PHG and ERC. These putatively adverse effects were observed regardless of genetic or CVD risks for ADRD and were detected even with low levels of exposures in compliance with regulatory standards. These findings support the role of air pollution neurotoxicity along the brain aging continuum. Future work includes investigating the underlying neuropathological processes and white matter microstructure affected by late-life exposures.

## Data Availability

Data, codebook, and analytic code used in this report are held by the NIH-funded Coordinating Center of the Women's Health Initiative at the Fred Hutchinson Cancer Research Center and may be accessed as described on the Women's Health Initiative website: https://www.whi.org/md/working-with-whi-data.

## Acknowledgements

The air pollution models were developed under a STAR research assistance agreement, No. RD831697 (MESA Air) and RD-83830001 (MESA Air Next Stage), awarded by the US Environmental Protection Agency (EPA). This study is supported by R01AG033078 (PI: Dr. Chen), RF1AG054068 (PI: Dr. Chen), R01ES025888 (PI: Drs. Chen & Kaufman), P01AG055367, the Southern California Environmental Health Sciences Center (5P30ES007048), and the Alzheimer’s Disease Research Center at USC (NIA; P50AG005142 and P30AG066530). Dr. Salminen is support by grant 1R01ES033961-01, which is co-funded by NIEHS and NIA. Dr. Younan is also supported by a grant from the Alzheimer’s Association (AARF-19-591356). Dr. Espeland receives funding from the Wake Forest Alzheimer’s Disease Core Center (P30AG049638–01A1). Dr. Resnick is supported by the Intramural Research Program, National Institute on Aging, NIH. The WHI program is funded by the National Heart, Lung, and Blood Institute, National Institutes of Health, U.S. Department of Health and Human Services through contracts HHSN268201600018C, HHSN268201600001C, HHSN268201600002C, HHSN268201600003C, and HHSN268201600004C. A list of contributors to WHI is available at https://www-whi-org.s3.us-west-2.amazonaws.com/wp-content/uploads/WHI-Investigator-Long-List.pdf.

## Role of the Funder/Sponsor

The funders/sponsors had no role in the design and conduct of the study; collection, management, analysis, and interpretation of the data; preparation, review, or approval of the manuscript; and decision to submit the manuscript for publication.

## Conflict of Interest Disclosures

None reported

## Author Contributions

Drs. Wang and Salminen contributed equally to this work.

Dr. Wang had full access to all the data in the study and takes responsibility for the integrity of the data and the accuracy of the data analysis.

Xinhui Wang: Conceptualization, Methodology, Software, Validataion, Formal analysis, Data Curation, Visualization, Writing - original draft;

Lauren E. Salminen: Conceptualization, Methodology, Writing - original draft;

Andrew J. Petkus: Methodology, Writing - Review & Editing;

Ira Driscoll: Conceptualization, Writing - Review & Editing;

Joshua Millstein: Methodology, Writing - Review & Editing;

Daniel P. Beavers: Methodology, Data Curation, Writing - Review & Editing;

Mark A. Espeland: Methodology, Data Curation, Writing - Review & Editing;

Guray Erus: Data Curation, Writing - Review & Editing;

Meredith N. Braskie: Conceptualization, Writing - Review & Editing;

Paul M. Thompson: Conceptualization, Writing - Review & Editing;

Margaret Gatz: Writing - Review & Editing;

Helena C. Chui: Writing - Review & Editing;

Susan M Resnick: Conceptualization, Writing - Review & Editing;

Joel D. Kaufman: Funding acquisition, Writing - Review & Editing;

Stephen R. Rapp: Conceptualization, Writing - Review & Editing;

Sally Shumaker: Conceptualization, Writing - Review & Editing;

Mark Brown: Data Curation, Writing - Review & Editing;

Diana Younan: Writing - Review & Editing;

Jiu-Chiuan Chen: Conceptualization, Supervision, Project administration, Funding acquisition, Writing - original draft

## Notes

### Competing Interest Statement

The authors have declared no competing interest.

