## Supplement for "Association between late-life air pollution exposure and medial temporal lobe atrophy in older women"

**Method S1.** Neighborhood socioeconomic status

**Method S2.** Assessment of covariates at MRI-1 visit

**Method S3.** APOE genotype data

**Method S4.** Inverse probability weighting approach to control for selective participation from MRI-1 to MRI-2

**Method S5.** Ascertainment of all-cause dementia in WHIMS

**Method S6.** *ad hoc* Cox proportional hazard regression

**Results of Table S3:** Sample characteristics associated with atrophy in MTL subregions

**Table S1.** Distribution of Continuous Measurements in the WHIMS MRI-1 Study Sample, Stratified by Women Included vs. Excluded due to missing data.

**Table S2.** Distribution of Population Characteristics in the WHIMS MRI-1 Study Sample, Stratified by Women Included vs. Excluded due to missing data.

**Table S3.** Distribution of standardized 5-year change in volumes of medial temporal lobe subregions by population characteristics.

**Table S4.** Associations between air pollution exposure and atrophy of medial temporal lobe subregions.

**Table S5.** Associations between PM_2.5_ exposure and atrophy of medial temporal lobe and its subregions, among women with PM_2.5_ less than NAAQS (12 µg/m^3^) (N=401).

**Table S6.** Associations between air pollution exposures and atrophy of medial temporal lobe and its subregions, excluding 33 women with incident MCI or dementia before MRI-2.

**Method S1. Neighborhood socioeconomic status**

Socioeconomic characteristics of residential neighborhood characterized at the U.S. Census tract-level were calculated at both WHI inception and MRI-1 visit (Diez Roux et al. 2001). Briefly, six attributes covering domains of wealth/income, education, and occupation from the U.S. Census of Population and Housing 2000 Summary File 3 (U.S. Census Bureau and Inter-university Consortium for Political and Social Research 2006), or their five-year analogs from American Community Survey 2005–2009 to 2013–2017 (U.S. Census Bureau), were temporally matched to geocoded participant addresses (Whitsel et al. 2004; Whitsel et al. 2006). Each attribute had been aggregated at the U.S. Census tract (i.e., the lowest geographic level historically associated with accurate and reliable assignment of Federal Information Processing System codes) (Whitsel et al. 2006). The 6 variables included: 1) log transformation of median household income; 2) log transformed median value of owner-occupied housing units; 3) percent of households receiving interest, dividend or net rent income; 4) percentage of adults ≥25 with a high school degree; 5) percentage of adults ≥25 with a college degree; and 6) percentage of employed persons aged ≥16 with a professional, managerial, or executive occupations. These variables were standardized using the corresponding population-specific mean and standard deviation, and then summed to derive a z-score for neighborhood socioeconomic status (SES). As computed, a higher neighborhood SES score implied a more advantageous neighborhood socioeconomic status.

**Method S2. Assessment of covariates at MRI-1 visit**

In addition to those covariates collected at the WHI inception, lifestyle factors, including smoking status (never, past or current smoker) and alcohol intake (grouped as non-drinker, past-drinker, <1 drink per day, or ≥1 drink per day), were updated using longitudinally collected smoking status (Yes vs. No), alcohol consumption (servings per week) before the MRI-1 visit. Physical activity (number of episodes per week of moderate and strenuous recreational physical activity of ≥20 minutes) was collected using a questionnaire administered at the closest visit before the MRI-1 visit. Body mass index was calculated using height and weight measured at the closest clinic visit before the MRI-1 visit. History of hypertension at the MRI-1 visit was defined as having a history of hypertension at WHI inception or having an elevated blood pressure (defined as systolic ≥140 or diastolic ≥90 mmHg) at any annual in-person visits before the MRI-1 visit. In addition to the self-reported history of cardiovascular disease (CVD) collected at the WHI inception, prospectively collected data on incident cardiovascular diseases (coronary heart disease; myocardial infarction; coronary revascularization; coronary angioplasty; coronary artery bypass graft; atrial fibrillation; stroke) were used to update the information on CVD histories prior to the MRI-1 visit. The binary indicator variable for CVD and related risk factors at MRI-1 was then updated using the updated hypertension and CVD history information.

**Method S3. APOE genotype data**

APOE genotypes were assigned based on rs429358 and rs7412 SNPs results, genotyped or imputed and harmonized across multiple WHI genome wide association studies. (Hayden et al. 2019) The imputation was conducted using the 1000 Genomes Project reference panel and the MaCH algorithm implemented in Minimac (R^2^ > 0.97 for both SNPs). (Howie et al. 2012) In analyses, women with APOE genotypes ε3/ε3 were treated as the reference group, while women with genotypes ε3/ε4 or ε4/ε4 were grouped together representing women with increased risk of AD.

**Method S4. Inverse probability weighting approach to control for selective participation from MRI-1 to MRI-2**

We implemented the inverse probability weighting approach (Hernan et al. 2004) in our analyses to control for possible bias due to selective participation in our study.

The weights were constructed using two steps. First, we used logistic regression to build a prediction model to determine an individual’s probability of participating in the longitudinal study ($Z_{i}=1$ if yes, 0 otherwise). We treated this indicator as the dependent variable. For predictor variables, we included all covariates (air pollution exposures; age; race/ethnicity; geographic region; socioeconomic factors; lifestyle factors; clinical characteristics; WHI-HT intervention assignment, and intracranial volume) in our model to determine which were significant predictors for being selected in the longitudinal analyses. Our final prediction model included air pollution exposure, geographic region, and income, which we denoted as covariate vector $X_{i}$. We then modeled the conditional probability of being selected using these predictor variables ($P(Z_{i}=z_{i}|X_{i}=x_{i})$) and output the estimated probabilities to calculate the crude weights ($cw_{i}$) as follows:

$${cw}_{i}=\frac{1}{P(Z_{i}=z_{i}|X_{i}=x_{i})}$$

Because we were conducting longitudinal analyses on those included participants ($Z_{i}=1$) (Seaman et al. 2013) only, their crude weights could also be written as:

$${cw}_{i}=\frac{1}{P(Z_{i}=1|X_{i}=x_{i})}$$

Next, we stabilized these weights to avoid unexpectedly large weights due to potential high missingness from subgroups. (Hernan et al. 2000; Weuve et al. 2012) This was done by running another logistic regression model using a reduced set of time-invariant covariates as predictors (race/ethnicity; WHI- HT intervention assignment), which we denoted as covariate vector $X_{i}'$. We modeled the conditional probability of participating in the longitudinal analyses ($P(Z_{i}=z_{i}|X_{i}'=x_{i}')$) and multiplied this by the crude weights generated above. Thus, for those included participants ($Z_{i}=1$), we calculated the stabilized weights (${sw}_{i}$) as follows:

$${sw}_{i}={cw}_{i}*P\left( Z_{i}=1 | X_{i}^{'}=x_{i}^{'} \right)=\frac{P(Z_{i}=1|X_{i}'=x_{i}')}{P(Z_{i}=1|X_{i}=x_{i})}$$

In our linear models regressing volumetric changes in medial temporal lobe on air pollution exposures and potential confounders, we specified a weight statement to account for the stabilized weights ${sw}_{i}$.

**Method S5. Ascertainment of all-cause dementia in WHIMS**

Incident cases of all-cause dementia were determined using published WHIMS protocols ( Shumaker et al., 2003; Shumaker et al. 2004; WHI Memory Study 2020). Briefly, in WHIMS on-trial and post-trial phases when participants had annual in-person visits, Modified Mini Mental State Exam (3MSE) was administered as cognitive function screening test. Later in the Epidemiology of Cognitive Health Outcomes (ECHO) phase since 2008, participants underwent annual validated telephone interview that used the modified Telephone Interview for Cognitive Status (TICSm) as the cognitive function screening test. If a participant scored ≤ the cut point (80 [or 72 before July 1, 1998] for women with ≤8 years of education and 88 [or 76 before July 1, 1998] for those with ≥9 years of education in the 3MSE test; 30 for the TICSm test), the standardized, validated Dementia Questionnaire (DQ) was administered to a previously identified proxy (friend or family member). The results and the cognitive scoring history were then reviewed by a panel of experts in the diagnosis of dementia. A Supplemental Case Ascertainment Protocol (SCAP) was also implemented to identify cases of dementia in the deceased and proxy-dependent participants (Gaussoin et al. 2019). In SCAP, the DQ data was also administrated to a participant-identified proxy and all prior assessments were used for adjudication of dementia. Clinical diagnoses of dementia were based on the Diagnostic and Statistical Manual of Mental Disorders (fourth edition) decided by the central adjudication committee for final confirmation.

**Method S6. *ad hoc* Cox proportional hazard regression**

To estimate how exposure-related MTL atrophy would translate to dementia risk, we conducted an *ad hoc* Cox proportional hazard regression to evaluate dementia risk associated with MTL volume among women without dementia before MRI-1, using outcome data from the WHIMS protocols. Follow-up time was defined as days since MRI-1 visit to the first occurrence of the cognitive assessment leading to the classification of dementia, death, or the last date of cognitive assessment (through June 2018), whichever came first. We incorporated inverse-probability weighting to account for differential attrition over follow up. Potential confounders included geographic region, sociodemographic characteristics, lifestyle factors, and clinical covariates at the WHI inception.

**Results of Table S3: Sample characteristics associated with atrophy in MTL subregions**

On average, the volumes of all 4 MTL subregions decreased over 5 years (Table S3). Women with BMI <25 kg/m^2^ at MRI-1 had greater atrophy in all 4 subregions, although the PHG atrophy did not reach statistical significance. Women ≥ 80 years of age or residing in the Northeast had greater atrophy in the PHG and hippocampus, but not in the ERC or amygdala. Greater PHG atrophy was also observed in women with higher income, resided in more socioeconomically-favorable neighborhoods at WHI inception or MRI-1, and those who consumed ≥1 alcoholic drink per day. Greater ERC atrophy was observed in women living in the Northeast and Midwest and those reported some physical activity (<2 episodes per week of moderate or strenuous physical activities that were ≥20 minutes). Women living in a more socioeconomically-favorable neighborhood at WHI inception or MRI-1 had less atrophy in the amygdala. Finally, women with at least one *APOE* ε4 allele had greater atrophy in the ERC and amygdala, but not the PHG or hippocampus, compared to *APOE* ε4 non-carriers (Table S3).

**Table S1. Distribution of Continuous Measurements in the WHIMS MRI-1 Study Sample, Stratified by Women Included vs. Excluded due to missing data.**

|  | **WHIMS MRI-1 study sample**  **(N=1405)^a^** | | **Analytic sample (N=653)** | | **Excluded due to missing data (N=752)^a^** | |  |
| --- | --- | --- | --- | --- | --- | --- | --- |
| **Measurements at MRI-1^b^** | **N** | **Mean ± SD** | **N** | **Mean ± SD** | **N** | **Mean ± SD** | **p^c^** |
| Age (years) | 1405 | 77.87 ± 3.70 | 653 | 77.34 ± 3.49 | 752 | 78.32 ± 3.81 | <0.001*** |
| PM_2.5_ exposure (µg/m^3^)^d^ | 1405 | 11.44 ± 2.48 | 653 | 11.19 ± 2.36 | 752 | 11.66 ± 2.56 | <0.001*** |
| NO_2_ exposure (ppb)^d^ | 1405 | 12.91 ± 6.77 | 653 | 11.92 ± 5.30 | 752 | 13.78 ± 7.73 | <0.001*** |
| Depressive symptoms at WHI inception^e^ | 1374 | 0.03 ± 0.09 | 653 | 0.03 ± 0.10 | 721 | 0.03 ± 0.09 | 0.91 |
| Neighborhood SES at WHI inception^f^ | 1351 | -0.03 ± 5.25 | 653 | 0.01 ± 5.14 | 698 | -0.06 ± 5.35 | 0.82 |
| Neighborhood SES at MRI-1^f^ | 1351 | -0.03 ± 5.25 | 653 | 0.01 ± 5.14 | 698 | -0.06 ± 5.35 | 0.82 |
| Total MTL Volume (cm^3^) | 1405 | 17.43 ± 1.79 | 653 | 17.76 ± 1.66 | 752 | 17.15 ± 1.85 | <0.001*** |
| Parahippocampal Gyrus Volume (cm^3^) | 1405 | 5.33 ± 0.65 | 653 | 5.41 ± 0.63 | 752 | 5.26 ± 0.65 | <0.001*** |
| Entorhinal Cortex Volume (cm^3^) | 1405 | 3.76 ± 0.58 | 653 | 3.85 ± 0.55 | 752 | 3.68 ± 0.59 | <0.001*** |
| Amygdala Volume (cm^3^) | 1405 | 1.76 ± 0.23 | 653 | 1.78 ± 0.21 | 752 | 1.73 ± 0.24 | <0.001*** |
| Hippocampus Volume (cm^3^) | 1405 | 6.58 ± 0.71 | 653 | 6.71 ± 0.65 | 752 | 6.47 ± 0.74 | <0.001*** |

Abbreviations: MRI, magnetic resonance imaging; SES, socioeconomic status; WHI, Women’s Health Initiative.

^a^ Numbers in the samples may not added up to total due to missing.

^b^ Measures were taken at the time of MRI-1 visit unless specified.

^c^ p-values were calculated using ANOVA F-tests for group comparisons. *p<0.05, **<0.01, ***<0.001

^d^ Air pollution exposures were preceding 3-year average estimated at the MRI-1 visit.

^e^ Depressive symptoms were assessed using the Center for Epidemiologic Studies Depression Scale short form. The score ranged from 0 to 1 with a higher score indicating a greater likelihood of depression.

^f^ Neighborhood SES is the sum of six standardized U.S. Census tract–level variables measuring domains of wealth/income, education, and occupation. A higher neighborhood SES score indicates a more advantageous neighborhood SES.

**Table S2. Distribution of Population Characteristics in the WHIMS MRI-1 Study Sample, Stratified by Women Included vs. Excluded due to missing data.**

| **Population Characteristics at WHI inception (1993-1998): N (%)** | **WHIMS MRI-1 study sample (N=1405)^a^** | **Analytic sample (N=653)** | **Excluded due to missing data (N=752)^a^** | **p^b^** |
| --- | --- | --- | --- | --- |
| **Sociodemographic Variables** |  |  |  |  |
| Age at MRI-1 visit (years) |  |  |  |  |
| < 75 | 344 (24.5%) | 189 (28.9%) | 155 (20.6%) | <0.001*** |
| ≥ 75 and < 80 | 662 (47.1%) | 314 (48.1%) | 348 (46.3%) |  |
| ≥ 80 | 399 (28.4%) | 150 (23.0%) | 249 (33.1%) |  |
| Region |  |  |  |  |
| Northeast | 330 (23.5%) | 172 (26.3%) | 158 (21.0%) | <0.001*** |
| South | 211 (15.0%) | 64 (9.8%) | 147 (19.5%) |  |
| Midwest | 489 (34.8%) | 255 (39.1%) | 234 (31.1%) |  |
| West | 375 (26.7%) | 162 (24.8%) | 213 (28.3%) |  |
| Race and Ethnicity |  |  |  |  |
| White (not Hispanic) | 1285 (91.5%) | 614 (94.0%) | 671 (89.3%) | 0.002** |
| Other ethnic or racial background^c^ | 119 (8.5%) | 39 (6.0%) | 80 (10.7%) |  |
| Education |  |  |  |  |
| ≤ High school or GED | 388 (27.7%) | 194 (29.7%) | 194 (25.9%) | 0.24 |
| > HS/GED but < 4y of college | 554 (39.5%) | 246 (37.7%) | 308 (41.1%) |  |
| ≥ 4y of college | 460 (32.8%) | 213 (32.6%) | 247 (33.0%) |  |
| Employment |  |  |  |  |
| Currently working | 174 (12.4%) | 85 (13.0%) | 89 (11.9%) | 0.51 |
| Not working | 119 (8.5%) | 60 (9.2%) | 59 (7.9%) |  |
| Retired | 1110 (79.1%) | 508 (77.8%) | 602 (80.3%) |  |
| Family Income |  |  |  |  |
| < $35,000 | 719 (51.2%) | 313 (47.9%) | 406 (54.0%) | 0.01* |
| $35,000 to $74,999 | 506 (36.0%) | 252 (38.6%) | 254 (33.8%) |  |
| ≥ $75,000 | 109 (7.8%) | 61 (9.3%) | 48 (6.4%) |  |
| Not known | 71 (5.1%) | 27 (4.1%) | 44 (5.9%) |  |
| **Lifestyle Factors** |  |  |  |  |
| Smoking status |  |  |  |  |
| Never smoked | 804 (57.7%) | 376 (57.6%) | 428 (57.8%) | 0.99 |
| Past smoker | 530 (38.0%) | 249 (38.1%) | 281 (38.0%) |  |
| Current Smoker | 59 (4.2%) | 28 (4.3%) | 31 (4.2%) |  |
| Alcohol use |  |  |  |  |
| Non-drinker | 181 (13.0%) | 82 (12.6%) | 99 (13.3%) | 0.91 |
| Past drinker | 232 (16.6%) | 105 (16.1%) | 127 (17.1%) |  |
| < 1 drink per day | 827 (59.3%) | 393 (60.2%) | 434 (58.5%) |  |
| ≥ 1 drink per day | 155 (11.1%) | 73 (11.2%) | 82 (11.1%) |  |
| Moderate or strenuous physical activities ≥ 20 minutes |  |  |  |  |
| No activity | 800 (57.0%) | 354 (54.2%) | 446 (59.5%) | 0.20 |
| Some activity | 79 (5.6%) | 40 (6.1%) | 39 (5.2%) |  |
| 2-4 episodes/week | 282 (20.1%) | 144 (22.1%) | 138 (18.4%) |  |
| > 4 episodes/week | 242 (17.2%) | 115 (17.6%) | 127 (16.9%) |  |
| **Physical Health** |  |  |  |  |
| Body Mass Index (kg/m^2^) |  |  |  |  |
| < 25 | 417 (29.8%) | 190 (29.1%) | 227 (30.4%) | 0.30 |
| 25-29 | 530 (37.9%) | 261 (40.0%) | 269 (36.0%) |  |
| ≥ 30 | 453 (32.4%) | 202 (30.9%) | 251 (33.6%) |  |
| CVD and related risk factors^e^ |  |  |  |  |
| None | 708 (50.4%) | 355 (54.4%) | 353 (46.9%) | 0.01* |
| At least 1 type | 697 (49.6%) | 298 (45.6%) | 399 (53.1%) |  |
| Any prior postmenopausal hormone use |  |  |  |  |
| No | 757 (53.9%) | 348 (53.3%) | 409 (54.4%) | 0.68 |
| Yes | 648 (46.1%) | 305 (46.7%) | 343 (45.6%) |  |
| WHI hormone therapy assignment |  |  |  |  |
| CEE-alone placebo | 265 (18.9%) | 116 (17.8%) | 149 (19.8%) | 0.72 |
| CEE-alone intervention | 258 (18.4%) | 118 (18.1%) | 140 (18.6%) |  |
| CEE+MPA placebo | 447 (31.8%) | 215 (32.9%) | 232 (30.9%) |  |
| CEE+MPA intervention | 435 (31.0%) | 204 (31.2%) | 231 (30.7%) |  |
| *APOE* genotype |  |  |  |  |
| ε3/3 | 860 (75.9%) | 402 (77.0%) | 458 (75.0%) | 0.42 |
| ε3/4+ε4/4 | 273 (24.1%) | 120 (23.0%) | 153 (25.0%) |  |
| **Covariates updated at MRI-1 (2005-2006)** |  |  |  |  |
| Smoking status |  |  |  |  |
| Never smoked | 793 (57.1%) | 371 (57.0%) | 422 (57.2%) | 0.98 |
| Past smoker | 552 (39.7%) | 260 (39.9%) | 292 (39.6%) |  |
| Current Smoker | 44 (3.2%) | 20 (3.1%) | 24 (3.3%) |  |
| Alcohol use |  |  |  |  |
| Non-drinker | 178 (12.8%) | 80 (12.3%) | 98 (13.2%) | 0.37 |
| Past drinker | 479 (34.3%) | 214 (32.8%) | 265 (35.7%) |  |
| < 1 drink per day | 607 (43.5%) | 300 (46.0%) | 307 (41.3%) |  |
| ≥ 1 drink per day | 131 (9.4%) | 58 (8.9%) | 73 (9.8%) |  |
| Moderate or strenuous physical activities ≥ 20 minutes |  |  |  |  |
| No activity | 819 (59.0%) | 377 (58.4%) | 442 (59.6%) | 0.78 |
| Some activity | 56 (4.0%) | 24 (3.7%) | 32 (4.3%) |  |
| 2-4 episodes/week | 290 (20.9%) | 142 (22.0%) | 148 (19.9%) |  |
| > 4 episodes/week | 222 (16.0%) | 102 (15.8%) | 120 (16.2%) |  |
| Body Mass Index (kg/m^2^) |  |  |  |  |
| < 25 | 428 (30.5%) | 185 (28.3%) | 243 (32.3%) | 0.01* |
| 25-29 | 528 (37.6%) | 272 (41.7%) | 256 (34.0%) |  |
| ≥ 30 | 449 (32.0%) | 196 (30.0%) | 253 (33.6%) |  |
| CVD and related risk factors^f^ |  |  |  |  |
| None | 289 (20.6%) | 148 (22.7%) | 141 (18.8%) | 0.07 |
| At least 1 type | 1116 (79.4%) | 505 (77.3%) | 611 (81.3%) |  |

Abbreviations: ApoE, Apolipoprotein E; CEE, conjugated equine estrogens; CVD, cardiovascular disease; HS/GED, high school or general educational development; MPA, medroxyprogesterone acetate; MRI, magnetic resonance imaging; SES, socioeconomic status; WHI, Women’s Health Initiative.

^a^ Numbers in the samples may not added up to total due to missing.

^b^ p-values were calculated using Chi-square tests. *p<0.05, **<0.01, ***<0.001

^C^ Others include Hispanic women or non-Hispanic women with race in American Indian/Alaska Native, Asian, Black, more than one race, or unknown/not reported.

^d^ Neighborhood SES is the sum of six standardized U.S. Census tract–level variables measuring domains of wealth/income, education, and occupation. A higher neighborhood SES score indicates a more advantageous neighborhood SES.

^e^ CVD and related risk factors were defined by whether a person had reported any history of CVD event (self-reported heart problems, problems with blood circulation, or blood clots), hypertension, hypercholesterolemia, or diabetes mellitus (none vs. at least one).

^f^ CVD and related risk factors were defined by whether a person had reported any history of CVD event (self-reported at WHI inception or any incident CVD events occurred before MRI-1), hypertension, hypercholesterolemia, or diabetes mellitus (none vs. at least one).

**Table S3. Distribution of standardized 5-year change in volumes of medial temporal lobe subregions by population characteristics.**

| **Population Characteristics at WHI** |  | **Atrophy in Parahippocampul Gyrus (cm^3^)** | | **Atrophy in Entorhinal Cortex (cm^3^)** | | **Atrophy in**  **Amygdala (cm^3^)** | | **Atrophy in Hippocampus (cm^3^)** | |
| --- | --- | --- | --- | --- | --- | --- | --- | --- | --- |
| **inception (1993-1998)** | **N (%)** | **Mean ± SD** | **p^a^** | **Mean ± SD** | **p^a^** | **Mean ± SD** | **p^a^** | **Mean ± SD** | **p^a^** |
| Overall | 653 (100%) | -0.04 ± 0.50 |  | -0.16 ± 0.41 |  | -0.08 ± 0.13 |  | -0.24 ± 0.33 |  |
| **Sociodemographic variables** |  |  |  |  |  |  |  |  |  |
| Age at MRI-1 visit (years) |  |  |  |  |  |  |  |  |  |
| < 75 | 189 (28.9%) | 0.03 ± 0.51 | 0.04* | -0.12 ± 0.42 | 0.24 | -0.07 ± 0.13 | 0.08 | -0.19 ± 0.39 | 0.007** |
| ≥ 75 and < 80 | 314 (48.1%) | -0.07 ± 0.50 |  | -0.17 ± 0.42 |  | -0.09 ± 0.13 |  | -0.24 ± 0.30 |  |
| ≥ 80 | 150 (23.0%) | -0.08 ± 0.46 |  | -0.19 ± 0.40 |  | -0.08 ± 0.13 |  | -0.30 ± 0.31 |  |
| Region |  |  |  |  |  |  |  |  |  |
| Northeast | 172 (26.3%) | -0.18 ± 0.33 | <0.001*** | -0.20 ± 0.34 | 0.003** | -0.09 ± 0.11 | 0.19 | -0.34 ± 0.32 | <0.001*** |
| South | 64 (9.8%) | 0.14 ± 0.57 |  | -0.06 ± 0.46 |  | -0.07 ± 0.19 |  | -0.20 ± 0.34 |  |
| Midwest | 255 (39.1%) | 0.00 ± 0.44 |  | -0.21 ± 0.41 |  | -0.09 ± 0.13 |  | -0.20 ± 0.34 |  |
| West | 162 (24.8%) | -0.05 ± 0.64 |  | -0.10 ± 0.45 |  | -0.07 ± 0.13 |  | -0.20 ± 0.32 |  |
| Race and Ethnicity |  |  |  |  |  |  |  |  |  |
| White (not Hispanic) | 614 (94.0%) | -0.04 ± 0.51 | 0.26 | -0.17 ± 0.41 | 0.18 | -0.08 ± 0.13 | 0.68 | -0.24 ± 0.33 | 0.88 |
| Other ethnic or racial background^b^ | 39 (6.0%) | -0.13 ± 0.33 |  | -0.08 ± 0.39 |  | -0.08 ± 0.12 |  | -0.23 ± 0.32 |  |
| Education |  |  |  |  |  |  |  |  |  |
| ≤ High school or GED | 194 (29.7%) | 0.00 ± 0.51 | 0.09 | -0.17 ± 0.40 | 0.83 | -0.09 ± 0.13 | 0.34 | -0.23 ± 0.35 | 0.52 |
| > HS/GED but < 4y of college | 246 (37.7%) | -0.03 ± 0.55 |  | -0.17 ± 0.41 |  | -0.08 ± 0.13 |  | -0.22 ± 0.33 |  |
| ≥ 4y of college | 213 (32.6%) | -0.10 ± 0.40 |  | -0.15 ± 0.43 |  | -0.07 ± 0.12 |  | -0.26 ± 0.32 |  |
| Employment |  |  |  |  |  |  |  |  |  |
| Currently working | 85 (13.0%) | 0.00 ± 0.56 | 0.11 | -0.16 ± 0.48 | 0.86 | -0.08 ± 0.12 | 0.84 | -0.20 ± 0.38 | 0.55 |
| Not working | 60 (9.2%) | -0.17 ± 0.40 |  | -0.14 ± 0.46 |  | -0.08 ± 0.15 |  | -0.22 ± 0.30 |  |
| Retired | 508 (77.8%) | -0.04 ± 0.49 |  | -0.17 ± 0.39 |  | -0.09 ± 0.13 |  | -0.24 ± 0.33 |  |
| Family Income |  |  |  |  |  |  |  |  |  |
| < $35,000 | 313 (47.9%) | 0.00 ± 0.53 | 0.01* | -0.19 ± 0.41 | 0.06 | -0.09 ± 0.13 | 0.76 | -0.23 ± 0.33 | 0.69 |
| $35,000 to $74,999 | 252 (38.6%) | -0.09 ± 0.45 |  | -0.15 ± 0.41 |  | -0.08 ± 0.12 |  | -0.24 ± 0.33 |  |
| ≥ $75,000 | 61 (9.3%) | -0.14 ± 0.41 |  | -0.18 ± 0.39 |  | -0.09 ± 0.13 |  | -0.27 ± 0.33 |  |
| Not known | 27 (4.1%) | 0.13 ± 0.57 |  | 0.03 ± 0.53 |  | -0.07 ± 0.14 |  | -0.18 ± 0.36 |  |
| Neighborhood SES^c^ |  |  |  |  |  |  |  |  |  |
| < -3.54 | 163 (25.0%) | 0.05 ± 0.54 | 0.01* | -0.17 ± 0.43 | 0.89 | -0.10 ± 0.14 | 0.002** | -0.25 ± 0.33 | 0.58 |
| ≥ -3.54 and < -0.26 | 162 (24.8%) | -0.04 ± 0.53 |  | -0.17 ± 0.42 |  | -0.10 ± 0.12 |  | -0.25 ± 0.35 |  |
| ≥ -0.26 and < 3.16 | 164 (25.1%) | -0.06 ± 0.44 |  | -0.17 ± 0.35 |  | -0.09 ± 0.13 |  | -0.25 ± 0.31 |  |
| ≥ 3.16 | 164 (25.1%) | -0.13 ± 0.45 |  | -0.14 ± 0.44 |  | -0.05 ± 0.12 |  | -0.21 ± 0.35 |  |
| **Lifestyle factors** |  |  |  |  |  |  |  |  |  |
| Smoking status |  |  |  |  |  |  |  |  |  |
| Never smoked | 376 (57.6%) | -0.06 ± 0.48 | 0.68 | -0.15 ± 0.42 | 0.49 | -0.08 ± 0.13 | 0.68 | -0.23 ± 0.31 | 0.59 |
| Past smoker | 249 (38.1%) | -0.03 ± 0.52 |  | -0.19 ± 0.40 |  | -0.08 ± 0.14 |  | -0.25 ± 0.36 |  |
| Current Smoker | 28 (4.3%) | 0.01 ± 0.54 |  | -0.11 ± 0.43 |  | -0.06 ± 0.12 |  | -0.19 ± 0.40 |  |
| Alcohol use |  |  |  |  |  |  |  |  |  |
| Non-drinker | 82 (12.6%) | -0.07 ± 0.43 | 0.03* | -0.17 ± 0.41 | 0.40 | -0.09 ± 0.15 | 0.43 | -0.25 ± 0.35 | 0.57 |
| Past drinker | 105 (16.1%) | 0.05 ± 0.60 |  | -0.11 ± 0.40 |  | -0.08 ± 0.13 |  | -0.21 ± 0.37 |  |
| < 1 drink per day | 393 (60.2%) | -0.04 ± 0.48 |  | -0.18 ± 0.41 |  | -0.09 ± 0.12 |  | -0.23 ± 0.31 |  |
| ≥ 1 drink per day | 73 (11.2%) | -0.18 ± 0.49 |  | -0.13 ± 0.43 |  | -0.06 ± 0.14 |  | -0.28 ± 0.38 |  |
| Moderate or strenuous physical activities ≥ 20 minutes | |  |  |  |  |  |  |  |  |
| No activity | 354 (54.2%) | -0.02 ± 0.52 | 0.40 | -0.16 ± 0.41 | 0.02* | -0.09 ± 0.13 | 0.41 | -0.25 ± 0.33 | 0.60 |
| Some activity | 40 (6.1%) | -0.15 ± 0.54 |  | -0.33 ± 0.28 |  | -0.08 ± 0.09 |  | -0.24 ± 0.32 |  |
| 2-4 episodes/week | 144 (22.1%) | -0.07 ± 0.44 |  | -0.11 ± 0.41 |  | -0.07 ± 0.12 |  | -0.23 ± 0.33 |  |
| > 4 episodes/week | 115 (17.6%) | -0.04 ± 0.48 |  | -0.20 ± 0.44 |  | -0.08 ± 0.15 |  | -0.20 ± 0.35 |  |
| **Physical Health** |  |  |  |  |  |  |  |  |  |
| Body Mass Index (kg/m^2^) |  |  |  |  |  |  |  |  |  |
| < 25 | 190 (29.1%) | -0.10 ± 0.50 | 0.12 | -0.18 ± 0.41 | 0.72 | -0.09 ± 0.13 | 0.51 | -0.25 ± 0.35 | 0.59 |
| 25-29 | 261 (40.0%) | 0.00 ± 0.49 |  | -0.16 ± 0.41 |  | -0.09 ± 0.13 |  | -0.24 ± 0.33 |  |
| ≥ 30 | 202 (30.9%) | -0.05 ± 0.50 |  | -0.15 ± 0.42 |  | -0.08 ± 0.13 |  | -0.22 ± 0.31 |  |
| CVD and related risk factors^d^ |  |  |  |  |  |  |  |  |  |
| None | 355 (54.4%) | -0.05 ± 0.51 | 0.60 | -0.17 ± 0.41 | 0.66 | -0.08 ± 0.13 | 0.39 | -0.25 ± 0.34 | 0.42 |
| At least 1 type | 298 (45.6%) | -0.03 ± 0.48 |  | -0.16 ± 0.41 |  | -0.09 ± 0.13 |  | -0.23 ± 0.33 |  |
| Any prior postmenopausal hormone use |  |  |  |  |  |  |  |  |  |
| No | 348 (53.3%) | -0.08 ± 0.47 | 0.07 | -0.19 ± 0.39 | 0.15 | -0.09 ± 0.12 | 0.14 | -0.24 ± 0.33 | 0.77 |
| Yes | 305 (46.7%) | -0.01 ± 0.52 |  | -0.14 ± 0.43 |  | -0.08 ± 0.14 |  | -0.23 ± 0.34 |  |
| WHI hormone therapy assignment |  |  |  |  |  |  |  |  |  |
| CEE-alone placebo | 116 (17.8%) | -0.01 ± 0.48 | 0.71 | -0.14 ± 0.42 | 0.65 | -0.08 ± 0.14 | 0.43 | -0.25 ± 0.38 | 0.58 |
| CEE-alone | 118 (18.1%) | -0.08 ± 0.52 |  | -0.19 ± 0.40 |  | -0.10 ± 0.13 |  | -0.27 ± 0.29 |  |
| CEE+MPA placebo | 215 (32.9%) | -0.05 ± 0.47 |  | -0.15 ± 0.42 |  | -0.08 ± 0.13 |  | -0.22 ± 0.33 |  |
| CEE+MPA | 204 (31.2%) | -0.04 ± 0.52 |  | -0.18 ± 0.40 |  | -0.08 ± 0.12 |  | -0.23 ± 0.33 |  |
| *APOE* genotype |  |  |  |  |  |  |  |  |  |
| ε3/3 | 402 (77.0%) | -0.06 ± 0.47 | 0.48 | -0.16 ± 0.40 | 0.05 | -0.07 ± 0.12 | 0.04* | -0.25 ± 0.32 | 0.77 |
| ε3/4+ε4/4 | 120 (23.0%) | -0.03 ± 0.52 |  | -0.24 ± 0.41 |  | -0.10 ± 0.15 |  | -0.24 ± 0.33 |  |
| **Covariates assessed at MRI-1 (2005-2006)** |  |  |  |  |  |  |  |  |  |
| Neighborhood SES^c^ |  |  |  |  |  |  |  |  |  |
| < -2.48 | 164 (25.1%) | 0.02 ± 0.50 | 0.002** | -0.20 ± 0.44 | 0.52 | -0.10 ± 0.14 | 0.048* | -0.25 ± 0.33 | 0.18 |
| ≥ -2.48 and < 0.67 | 163 (25.0%) | 0.02 ± 0.59 |  | -0.13 ± 0.41 |  | -0.09 ± 0.12 |  | -0.19 ± 0.34 |  |
| ≥ 0.67 and < 4.19 | 163 (25.0%) | -0.04 ± 0.43 |  | -0.17 ± 0.36 |  | -0.08 ± 0.12 |  | -0.27 ± 0.31 |  |
| ≥ 4.19 | 163 (25.0%) | -0.17 ± 0.42 |  | -0.16 ± 0.43 |  | -0.06 ± 0.14 |  | -0.23 ± 0.35 |  |
| Smoking status |  |  |  |  |  |  |  |  |  |
| Never smoked | 371 (57.0%) | -0.05 ± 0.48 | 0.43 | -0.15 ± 0.42 | 0.76 | -0.08 ± 0.13 | 0.77 | -0.23 ± 0.31 | 0.61 |
| Past smoker | 260 (39.9%) | -0.02 ± 0.52 |  | -0.18 ± 0.40 |  | -0.08 ± 0.14 |  | -0.24 ± 0.37 |  |
| Current Smoker | 20 (3.1%) | -0.16 ± 0.52 |  | -0.18 ± 0.42 |  | -0.06 ± 0.12 |  | -0.30 ± 0.36 |  |
| Alcohol use |  |  |  |  |  |  |  |  |  |
| Non-drinker | 80 (12.3%) | -0.06 ± 0.43 | 0.14 | -0.17 ± 0.41 | 0.89 | -0.09 ± 0.15 | 0.33 | -0.24 ± 0.35 | 0.93 |
| Past drinker | 214 (32.8%) | 0.01 ± 0.54 |  | -0.17 ± 0.39 |  | -0.08 ± 0.12 |  | -0.22 ± 0.33 |  |
| < 1 drink per day | 300 (46.0%) | -0.06 ± 0.50 |  | -0.16 ± 0.43 |  | -0.09 ± 0.13 |  | -0.24 ± 0.32 |  |
| ≥ 1 drink per day | 58 (8.9%) | -0.15 ± 0.39 |  | -0.12 ± 0.42 |  | -0.05 ± 0.14 |  | -0.23 ± 0.40 |  |
| Moderate or strenuous physical activities ≥ 20 minutes |  |  |  |  |  |  |  |  |  |
| No activity | 377 (58.4%) | -0.05 ± 0.51 | 0.79 | -0.20 ± 0.41 | 0.11 | -0.09 ± 0.14 | 0.16 | -0.25 ± 0.35 | 0.81 |
| Some activity | 24 (3.7%) | 0.00 ± 0.48 |  | -0.17 ± 0.40 |  | -0.08 ± 0.12 |  | -0.21 ± 0.28 |  |
| 2-4 episodes/week | 142 (22.0%) | -0.07 ± 0.42 |  | -0.10 ± 0.42 |  | -0.06 ± 0.13 |  | -0.23 ± 0.33 |  |
| > 4 episodes/week | 102 (15.8%) | -0.01 ± 0.52 |  | -0.14 ± 0.41 |  | -0.08 ± 0.11 |  | -0.22 ± 0.29 |  |
| Body Mass Index (kg/m^2^) |  |  |  |  |  |  |  |  |  |
| < 25 | 185 (28.3%) | -0.11 ± 0.50 | 0.08 | -0.23 ± 0.42 | 0.049* | -0.10 ± 0.14 | 0.02* | -0.30 ± 0.33 | 0.01* |
| 25-29 | 272 (41.7%) | -0.01 ± 0.49 |  | -0.14 ± 0.40 |  | -0.09 ± 0.12 |  | -0.23 ± 0.35 |  |
| ≥ 30 | 196 (30.0%) | -0.02 ± 0.50 |  | -0.13 ± 0.42 |  | -0.06 ± 0.13 |  | -0.19 ± 0.31 |  |
| CVD and related risk factors^e^ |  |  |  |  |  |  |  |  |  |
| None | 148 (22.7%) | -0.04 ± 0.51 | 0.97 | -0.15 ± 0.40 | 0.71 | -0.08 ± 0.13 | 0.90 | -0.22 ± 0.35 | 0.54 |
| At least 1 type | 505 (77.3%) | -0.04 ± 0.49 |  | -0.17 ± 0.42 |  | -0.08 ± 0.13 |  | -0.24 ± 0.33 |  |

Abbreviations: ApoE, Apolipoprotein E; CEE, conjugated equine estrogens; CVD, cardiovascular disease; GED, general educational development; MPA, medroxyprogesterone acetate; MRI, magnetic resonance imaging; MTL, medial temporal lobe; SD, standard deviation; SES, socioeconomic status; WHI, Women’s Health Initiative.

^a^ p-values were calculated using ANOVA F-tests for mean exposures. *p<0.05, **<0.01, ***<0.001

^b^ Others include Hispanic women or non-Hispanic women with race in American Indian/Alaska Native, Asian, Black, more than one race, or unknown/not reported.

^c^ Neighborhood SES is the sum of six standardized U.S. Census tract–level variables measuring domains of wealth/income, education, and occupation. A higher neighborhood SES score indicates a more advantageous neighborhood SES.

^d^ CVD and related risk factors were defined by whether a person had reported any history of CVD event (self-reported heart problems, problems with blood circulation, or blood clots), hypertension, hypercholesterolemia, or diabetes mellitus (none vs. at least one).

^e^ CVD and related risk factors were defined by whether a person had reported any history of CVD event (self-reported at WHI inception or any incident CVD events occurred before MRI-1), hypertension, hypercholesterolemia, or diabetes mellitus (none vs. at least one).

**Table S4. Associations between air pollution exposure and atrophy of medial temporal lobe subregions.**

a) **PM_2.5_ exposure^a^**

|  | **Parahippocampul Gyrus** | | | **Entorhinal Cortex** | | | **Amygdala** | | | **Hippocampus** | | |
| --- | --- | --- | --- | --- | --- | --- | --- | --- | --- | --- | --- | --- |
| **Models** | **b^b^** | **95% CI** | **p^c^** | **b^b^** | **95% CI** | **p^c^** | **b^b^** | **95% CI** | **p^c^** | **b^b^** | **95% CI** | **p^c^** |
| Model A | -0.23 | -0.28, -0.17 | <0.01 | -0.06 | -0.10, -0.01 | 0.03 | -0.005 | -0.02, 0.01 | 0.53 | -0.03 | -0.07, 0.004 | 0.11 |
| Model B | -0.24 | -0.29, -0.19 | <0.01 | -0.05 | -0.10, -0.004 | 0.07 | -0.001 | -0.02, 0.01 | 0.94 | -0.02 | -0.06, 0.02 | 0.33 |
| Model C | -0.24 | -0.30, -0.19 | <0.01 | -0.06 | -0.10, -0.01 | 0.04 | -0.001 | -0.02, 0.01 | 0.90 | -0.02 | -0.06, 0.02 | 0.37 |
| Model D | -0.24 | -0.29, -0.19 | <0.01 | -0.06 | -0.10, -0.01 | 0.04 | -0.001 | -0.02, 0.01 | 0.93 | -0.02 | -0.06, 0.02 | 0.36 |
| Model E | -0.24 | -0.29, -0.19 | <0.01 | -0.06 | -0.10, -0.01 | 0.04 | -0.001 | -0.02, 0.01 | 0.93 | -0.02 | -0.06, 0.02 | 0.36 |
| Model F | -0.23 | -0.29, -0.18 | <0.01 | -0.05 | -0.10, -0.01 | 0.06 | -0.002 | -0.02, 0.01 | 0.80 | -0.03 | -0.06, 0.01 | 0.25 |
| Model G | -0.24 | -0.29, -0.185 | <0.01 | -0.06 | -0.10, -0.01 | 0.03 | -0.002 | -0.02, 0.01 | 0.79 | -0.02 | -0.06, 0.02 | 0.40 |

b) **NO_2_ exposure^a^**

|  | **Parahippocampul Gyrus** | | | **Entorhinal Cortex** | | | **Amygdala** | | | **Hippocampus** | | |
| --- | --- | --- | --- | --- | --- | --- | --- | --- | --- | --- | --- | --- |
| **Models** | **b^b^** | **95% CI** | **p^c^** | **b^b^** | **95% CI** | **p^c^** | **b^b^** | **95% CI** | **p^c^** | **b^b^** | **95% CI** | **p^c^** |
| Model A | -0.12 | -0.16, -0.07 | <0.01 | -0.03 | -0.06, 0.01 | 0.34 | 0.000 | -0.01, 0.01 | >0.99 | -0.02 | -0.05, 0.01 | 0.34 |
| Model B | -0.08 | -0.13, -0.03 | <0.01 | -0.03 | -0.07, 0.02 | 0.45 | -0.002 | -0.02, 0.01 | 0.81 | -0.004 | -0.04, 0.03 | 0.81 |
| Model C | -0.09 | -0.14, -0.04 | <0.01 | -0.02 | -0.07, 0.02 | 0.68 | -0.002 | -0.02, 0.01 | 0.90 | -0.002 | -0.04, 0.03 | 0.90 |
| Model D | -0.09 | -0.14, -0.04 | <0.01 | -0.02 | -0.06, 0.02 | 0.73 | -0.001 | -0.02, 0.01 | 0.89 | -0.004 | -0.04, 0.03 | 0.89 |
| Model E | -0.09 | -0.14, -0.04 | <0.01 | -0.02 | -0.07, 0.02 | 0.69 | -0.001 | -0.02, 0.01 | 0.91 | -0.01 | -0.04, 0.03 | 0.91 |
| Model F | -0.09 | -0.14, -0.03 | <0.01 | -0.02 | -0.07, 0.02 | 0.72 | -0.001 | -0.02, 0.01 | 0.97 | -0.001 | -0.04, 0.03 | 0.97 |
| Model G | -0.09 | -0.14, -0.04 | <0.01 | -0.03 | -0.07, 0.02 | 0.51 | -0.004 | -0.02, 0.01 | 0.78 | -0.01 | -0.04, 0.03 | 0.78 |

Abbreviations: MRI, magnetic resonance imaging; MTL, medial temporal lobe; NO_2_, nitrogen dioxide; PM_2.5_, fine particulate matter; ppb, parts per billion; SES, socioeconomic status; WHI, Women’s Health Initiative.

^a^ Air pollution exposures were preceding 3-year average estimated at the MRI-1 visit.

interquartile range (IQR)_PM2.5_ = 3.26 µg/m^3^; IQR_NO2_ = 6.77 ppb

^b^ b represents the average change in brain volume (cm^3^) over 5 years for each IQR increase of 3-year average exposure. A negative b means higher air pollution exposure was associated with a greater atrophy in brain volume over 5 years.

Model A: adjusted for the intracranial volume and age

Model B: adjusted for Model A covariates + geographic region, race/ethnicity, education, income, employment status, and neighborhood SES

Model C: adjusted for Model B covariates + lifestyle factors (smoking; alcohol use; physical activities), prior postmenopausal hormone use, and WHI hormone therapy assignment

Model D: adjusted for Model C covariates + body mass index and depressive symptoms (logit transformation of the raw score)

Model E: adjusted for Model D covariates + cardiovascular disease and related risk factors

Model F: adjusted for Model E covariates with lifestyle factors, body mass index, neighborhood SES, and cardiovascular disease and related risk factors updated at MRI-1 visit.

Model G: adjusted for Model E covariates + corresponding brain volume at MRI-1 visit.

^c^ P values were corrected using Benjamini–Hochberg false discovery rate method across 4 MTL subregions for each model respectively.

**Table S5. Associations between PM_2.5_ exposure and atrophy of medial temporal lobe and its subregions, among women with PM_2.5_ less than NAAQS (12 µg/m^3^) (N=401).**

|  | **Total MTL** | | | **Parahippocampul Gyrus** | | | **Entorhinal Cortex** | | | **Amygdala** | | | **Hippocampus** | | |
| --- | --- | --- | --- | --- | --- | --- | --- | --- | --- | --- | --- | --- | --- | --- | --- |
| **Models** | **b^b^** | **95% CI** | **p** | **b^b^** | **95% CI** | **p^c^** | **b^b^** | **95% CI** | **p^c^** | **b^b^** | **95% CI** | **p^c^** | **b^b^** | **95% CI** | **p^c^** |
| Model A | -0.43 | -0.63, -0.22 | <0.01 | -0.33 | -0.44, -0.22 | <0.01 | -0.11 | -0.19, -0.02 | 0.03 | 0.001 | -0.03, 0.03 | 0.93 | 0.005 | -0.06, 0.07 | 0.93 |
| Model B | -0.44 | -0.65, -0.23 | <0.01 | -0.33 | -0.44, -0.22 | <0.01 | -0.10 | -0.19, -0.01 | 0.048 | -0.004 | -0.03, 0.03 | 0.88 | -0.01 | -0.07, 0.06 | 0.88 |
| Model C | -0.39 | -0.60, -0.18 | <0.01 | -0.30 | -0.41, -0.20 | <0.01 | -0.09 | -0.18, 0.001 | 0.11 | -0.003 | -0.03, 0.03 | 0.86 | 0.01 | -0.06, 0.08 | 0.86 |
| Model D | -0.39 | -0.60, -0.17 | <0.01 | -0.30 | -0.41, -0.19 | <0.01 | -0.09 | -0.18, 0.002 | 0.11 | -0.003 | -0.03, 0.03 | 0.91 | 0.004 | -0.07, 0.07 | 0.91 |
| Model E | -0.39 | -0.60, -0.18 | <0.01 | -0.30 | -0.41, -0.19 | <0.01 | -0.09 | -0.18, 0.001 | 0.11 | -0.003 | -0.03, 0.03 | 0.92 | 0.003 | -0.07, 0.07 | 0.92 |

Abbreviations: MRI, magnetic resonance imaging; MTL, medial temporal lobe; PM_2.5_, fine particulate matter; WHI, Women’s Health Initiative.

^a^ Air pollution exposures were 3-year average exposures estimated at the MRI-1 visit.

interquartile range (IQR)_PM2.5_ = 3.26 µg/m^3^

^b^ b represents the average change in brain volume (cm^3^) over 5 years for each IQR increase of 3-year average exposure. A negative b means higher air pollution exposure was associated with a greater atrophy in brain volume over 5 years.

Model A: adjusted for the intracranial volume and age

Model B: adjusted for Model A covariates + geographic region, race/ethnicity, education, income, employment status, and neighborhood socioeconomic status

Model C: adjusted for Model B covariates + lifestyle factors (smoking; alcohol use; physical activities), prior postmenopausal hormone use, and WHI hormone therapy assignment

Model D: adjusted for Model C covariates + body mass index and depressive symptoms (logit transformation of the raw score)

Model E: adjusted for Model D covariates + cardiovascular disease and related risk factors

^c^ P values were corrected using Benjamini–Hochberg false discovery rate method across 4 MTL subregions for each model respectively.

**Table S6. Associations between air pollution exposures and atrophy of medial temporal lobe and its subregions, excluding 33 women with incident MCI or dementia before MRI-2.**

a) **PM_2.5_ exposure^a^**

|  | **Total MTL** | | | **Parahippocampul Gyrus** | | | **Entorhinal Cortex** | | | **Amygdala** | | | **Hippocampus** | | |
| --- | --- | --- | --- | --- | --- | --- | --- | --- | --- | --- | --- | --- | --- | --- | --- |
| **Models** | **b^b^** | **95% CI** | **p** | **b^b^** | **95% CI** | **p^c^** | **b^b^** | **95% CI** | **p^c^** | **b^b^** | **95% CI** | **p^c^** | **b^b^** | **95% CI** | **p^c^** |
| Model A | -0.31 | -0.42, -0.21 | <0.01 | -0.22 | -0.28, -0.17 | <0.01 | -0.05 | -0.10, -0.01 | 0.04 | -0.005 | -0.02, 0.01 | 0.50 | -0.03 | -0.07, 0.01 | 0.13 |
| Model B | -0.31 | -0.42, -0.20 | <0.01 | -0.24 | -0.29, -0.19 | <0.01 | -0.05 | -0.09, -0.00 | 0.10 | -0.001 | -0.02, 0.01 | 0.92 | -0.02 | -0.06, 0.02 | 0.40 |
| Model C | -0.32 | -0.42, -0.21 | <0.01 | -0.24 | -0.30, -0.19 | <0.01 | -0.05 | -0.10, -0.01 | 0.05 | -0.001 | -0.02, 0.01 | 0.89 | -0.02 | -0.06, 0.02 | 0.42 |
| Model D | -0.32 | -0.42, -0.21 | <0.01 | -0.24 | -0.29, -0.19 | <0.01 | -0.05 | -0.10, -0.01 | 0.06 | -0.001 | -0.02, 0.01 | 0.91 | -0.02 | -0.06, 0.02 | 0.41 |
| Model E | -0.31 | -0.42, -0.21 | <0.01 | -0.24 | -0.29, -0.19 | <0.01 | -0.05 | -0.10, -0.01 | 0.06 | -0.001 | -0.02, 0.01 | 0.91 | -0.02 | -0.06, 0.02 | 0.41 |

b) **NO_2_ exposure^a^**

|  | **Total MTL** | | | **Parahippocampul Gyrus** | | | **Entorhinal Cortex** | | | **Amygdala** | | | **Hippocampus** | | |
| --- | --- | --- | --- | --- | --- | --- | --- | --- | --- | --- | --- | --- | --- | --- | --- |
| **Models** | **b^b^** | **95% CI** | **p** | **b^b^** | **95% CI** | **p^c^** | **b^b^** | **95% CI** | **p^c^** | **b^b^** | **95% CI** | **p^c^** | **b^b^** | **95% CI** | **p^c^** |
| Model A | -0.18 | -0.27, -0.09 | <0.01 | -0.13 | -0.18, -0.09 | <0.01 | -0.03 | -0.07, 0.01 | 0.24 | 0.001 | -0.01, 0.01 | 0.93 | -0.02 | -0.05, 0.02 | 0.46 |
| Model B | -0.13 | -0.23, -0.03 | 0.01 | -0.09 | -0.14, -0.04 | <0.01 | -0.04 | -0.08, 0.01 | 0.20 | -0.003 | -0.02, 0.01 | 0.87 | -0.003 | -0.04, 0.03 | 0.87 |
| Model C | -0.14 | -0.24, -0.04 | <0.01 | -0.11 | -0.16, -0.05 | <0.01 | -0.03 | -0.08, 0.01 | 0.33 | -0.002 | -0.02, 0.01 | 0.99 | 0.000 | -0.04, 0.04 | 0.99 |
| Model D | -0.14 | -0.24, -0.03 | <0.01 | -0.11 | -0.16, -0.05 | <0.01 | -0.03 | -0.07, 0.01 | 0.36 | -0.002 | -0.02, 0.01 | 0.93 | -0.002 | -0.04, 0.03 | 0.93 |
| Model E | -0.14 | -0.25, -0.04 | <0.01 | -0.11 | -0.16, -0.05 | <0.01 | -0.03 | -0.08, 0.01 | 0.34 | -0.002 | -0.02, 0.01 | 0.88 | -0.003 | -0.04, 0.03 | 0.88 |

Abbreviations: MRI, magnetic resonance imaging; MTL, medial temporal lobe; NO_2_, nitrogen dioxide; PM_2.5_, fine particulate matter; ppb, parts per billion; WHI, Women’s Health Initiative.

^a^ Air pollution exposures were 3-year average exposures estimated at the MRI-1 visit.

interquartile range (IQR)_PM2.5_ = 3.26 µg/m^3^; IQR_NO2_ = 6.77 ppb

^b^ b represents the average change in brain volume (cm^3^) over 5 years for each IQR increase of 3-year average exposure. A negative b means higher air pollution exposure was associated with a greater atrophy in brain volume over 5 years.

Model A: adjusted for the intracranial volume and age

Model B: adjusted for Model A covariates + geographic region, race/ethnicity, education, income, employment status, and neighborhood socioeconomic status

Model C: adjusted for Model B covariates + lifestyle factors (smoking; alcohol use; physical activities), prior postmenopausal hormone use, and WHI hormone therapy assignment

Model D: adjusted for Model C covariates + body mass index and depressive symptoms (logit transformation of the raw score)

Model E: adjusted for Model D covariates + cardiovascular disease and related risk factors

^c^ P values were corrected using Benjamini–Hochberg false discovery rate method across 4 MTL subregions for each model respectively.
